# A Multi-stage Precision Stratification (MPS) Framework for Navigating Adjuvant Immunotherapy in Hepatocellular Carcinoma After Resection

**DOI:** 10.64898/2026.08.08.26360002

**Authors:** Zhongfeng Dang, Jianduojie Dan, Wei Su, Guoliang Ren, Zhiqiang Wang, Yabing Ma, Shengmei Li, Dongde Ji, Liansheng Li, Junlin Gao, Yamei Dang

**Author notes:** **Co-first Authors:** Z. Dang and J. Dan contributed equally to this article. **Co-first Author Contact:** **Jianduojie Dan**, Department of Software Engineering, American University of Central Asia, Bishkek 720060, Kyrgyzstan. Primary Corresponding Author: **Zhongfeng Dang**, Department of Hepatobiliary and Pancreatic Surgery, Qinghai Red Cross Hospital, 55 Nandajie Street, Chengzhong District, Xining 810000, Qinghai, China. (Co-corresponding Authors:) **Junlin Gao‡**, Department of Hepatobiliary and Pancreatic Surgery, Qinghai Red Cross Hospital, Xining 810000, China.; **Yamei Dang‡**, Department of Pathology, Gansu Provincial People’s Hospital, Lanzhou 730000, China.

## Abstract

**Background:** Recurrence rates following curative resection for hepatocellular carcinoma (HCC) remain persistently high, benefit from adjuvant immunotherapy varies substantially across patients, and the field currently lacks a standardized framework to characterize the postoperative host immune contexture.

**Purpose:** To propose and validate a Multi-stage Precision Stratification (MPS) framework and evaluate its value in prognostic stratification and prediction of immunotherapy response.

**Methods:** The Immune Health Index (IHI = S + R − E) integrating immune surveillance (S), immune exhaustion (E), and immune reserve (R) was constructed to define four immune phenotypes. Prognostic value was assessed in four public HCC cohorts (n=931) with single-cell transcriptomic validation (GSE140228, 61,690 cells); a blood-count-based clinical version cIHI_v8 was constructed in the Qinghai QPHCC cohort (n=490 survival analysis).

**Results:** IHI was an independent protective prognostic factor in TCGA-LIHC (multivariate HR=0.795, P=0.034); four-cohort random-effects meta-analysis yielded HR=0.818 (95% CI: 0.696–0.961), I^2^=31.4%. QPHCC cIHI_v8 multivariate HR=0.452, HR=0.715 after ALBI adjustment; Bayesian evidence synthesis yielded BF_10=1280 for cIHI_v8 (>100 constitutes Decisive evidence), whereas the 4-cohort meta BF_10=2.19 (Anecdotal). Following NLP-based reverse stage derivation (n=490, achieving full AJCC/BCLC stage coverage from 0%), IHI remained significant after AJCC adjustment (HR=0.8642, P=0.000079), IHI provided positive incremental C-index across all stage-adjusted models; stratified analysis showed the strongest effect in early-stage (AJCC I-II: HR=0.8109, P<0.0001) and MVI-negative patients (HR=0.8538, P=0.0020). Bootstrap 1000× resampling: median HR=0.8646 (95% CI: 0.7985–0.9443), all iterations yielded HR<1.

**Conclusions:** The MPS framework provides a mechanism-driven biological stratification tool for adjuvant immunotherapy in post-resection HCC, moving from “fixed-protocol extrapolation” to “immune contexture navigation.”

## Introduction

Hepatocellular carcinoma (HCC) is a leading cause of cancer-related mortality worldwide[1]. For patients with resectable disease, curative-intent surgery offers the opportunity for long-term survival; however, the 5-year postoperative recurrence rate remains as high as 50–70%[2], substantially limiting the curative potential of surgery. Identifying high-risk patients and optimizing postoperative adjuvant therapy has remained a central challenge in HCC management.

In recent years, systemic therapies based on immune checkpoint inhibitors (ICIs) have been advanced into the adjuvant setting. The IMbrave050 trial[3] first reported positive results for atezolizumab combined with bevacizumab (T+A) as adjuvant therapy for high-risk HCC after resection: the 12-month recurrence-free survival (RFS) rate was 78% vs 65% (HR=0.55), yet with extended follow-up the survival curves gradually converged (updated analysis HR converged to approximately 0.76) and the long-term benefit was not sustained. Meanwhile, the KEYNOTE-937 trial[4] evaluating adjuvant pembrolizumab monotherapy yielded entirely negative results—median RFS 46.7 vs 45.5 months, with no statistical difference. These seemingly contradictory results raise a core question: why does the same immunotherapy produce diametrically opposite outcomes across different trials, and even among different patients within the same trial?

Analyzing the mechanistic differences between these trials provides clues. The “early-positive” phenomenon of IMbrave050 suggests that early postoperative micrometastases are highly dependent on the PD-L1/VEGF axis for immune evasion, and T+A can effectively eradicate these minimal residual lesions through dual PD-L1 blockade and VEGF inhibition; however, the “late-negative” phenomenon indicates that late recurrences beyond 18 months primarily arise from de novo tumors driven by chronic liver disease, whose immune evasion does not depend on the PD-L1 axis[5]. The failure of KEYNOTE-937 illustrates that in the complex postoperative microenvironment, PD-1 blockade alone is far from sufficient—the postoperative liver is caught in a “triple predicament”: immune exhaustion (terminal exhaustion of CD8^+^T cells), fibrotic barrier (extracellular matrix physically obstructing immune infiltration), and regeneration-driven tumor promotion (postoperative liver regeneration releasing HGF, TNF-α, and IL-6 that activate residual tumor stemness). By contrast, the ALTER-H006 trial[6] achieved positive results in a high-risk population by simultaneously covering “soil amendment” and “seed remodeling” (detailed in Discussion §1). These observations suggest that the efficacy of adjuvant immunotherapy depends critically on the host immune contexture at the time of intervention.

These successes and failures collectively point to a core insight: adjuvant immunotherapy can be effective, but only in specific populations and during specific time windows. What the field lacks is not more potent drugs, but a standardized framework capable of characterizing the postoperative host immune contexture and guiding stratified intervention. The wound-healing response triggered by surgery evolved to promote tissue repair, yet this same reparative process is accompanied by a transient but clinically meaningful immunosuppressive state—surgical trauma and intraoperative blood loss reduce effector T-cell and NK-cell counts while expanding Treg and myeloid-derived suppressor populations, creating a window of vulnerability during which residual micrometastases may evade immune surveillance[7–9]. The postoperative period therefore represents a unique “microenvironmental remodeling window”—the central question of adjuvant immunotherapy is no longer “which drug is more potent,” but rather “what immune contexture is the patient currently in, and which intervention mechanism best fits that contexture.” Accordingly, we propose the Multi-stage Precision Stratification (MPS) framework, aiming to shift the research logic of postoperative adjuvant immunotherapy from “empirical trial-and-error” to “immune contexture navigation.”

## Methods

### Study Design

The core of this study lies in proposing the Multi-stage Precision Stratification (MPS) as a conceptual framework, with the Immune Health Index (IHI) serving as the quantitative mathematical tool for implementing this framework. Based on the score combinations of IHI and its sub-modules, the MPS framework is operationalized into four actionable immune phenotypes. The study comprises two tiers: (1) bulk transcriptomic cohort analysis to evaluate the association of MPS with clinical outcomes; and (2) single-cell transcriptomic re-analysis to examine the cellular-level basis of key MPS phenotypes.

### Public Cohorts

Four independent public HCC bulk transcriptomic cohorts were included for validation: TCGA-LIHC (n=363), ICGC-LIRI-JP (n=232), GSE14520 (n=221), and GSE76427 (n=115). All cohorts were used as validation sets; the IHI formula was calculated independently within each cohort, with no cross-cohort training. Inclusion criteria were: (1) pathologically confirmed HCC; (2) availability of gene expression matrices; and (3) availability of survival outcome data. Exclusion criteria were: missing or unmappable expression data, duplicate samples, and missing clinical outcome information.

### Construction of the Immune Health Index (IHI)

IHI integrates three dimensions:

- Immune Surveillance module (S): CD8A, PRF1, GZMB, IFNG, CXCL9 — representing effector anti-tumor immune activity
- Immune Exhaustion module (E): PDCD1, LAG3, HAVCR2, TIGIT, TOX — representing T-cell exhaustion and inhibitory immune networks
- Immune Reserve module (R): TCF7, IL7R, CCR7, SELL, LTB — representing immune maintenance and mobilizable potential

Within each cohort, the expression value of each gene was standardized to a z-score. Module scores were calculated as the mean of the z-scores of genes within each module. IHI was defined as:

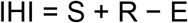

The linear additive structure of IHI is based on fundamental principles of tumor immunology: S and R represent complementary positive immune vectors, while E acts as a negative vector that offsets their function. To avoid high-variance weight estimation in small samples, an equal-weight linear combination was adopted, drawing on the equal-weight philosophy of Ayers TIS[10] and Galon Immunoscore[11]—under limited sample sizes, equal-weight combinations exhibit superior generalizability compared to empirical weight fitting. The variance comparison of the three modules’ relative contributions is provided in the Supplementary Materials.

### MPS Immune Phenotype Allocation

Based on IHI and its component modules, a rule-based approach (using the within-cohort median of each component as the high/low cutpoint) was used to classify patients into four immune phenotypes:

- Immune-activated phenotype: high IHI, low E — relatively preserved immune surveillance and effector function
- Immune-exhausted phenotype: high E and high S — effector immune components present but constrained by persistent inhibition
- Immune-fragile phenotype: low R and low S — limited immune reserve and weak effector function
- Immune-homeostatic phenotype: remaining patients — intermediate or relatively balanced immune status

Phenotype allocation follows the above order with priority matching: first identify the activated phenotype (IHI above median and E below median), then the exhausted phenotype (both E and S above median), then the fragile phenotype (both R and S below median); patients not meeting any of these criteria are assigned to the homeostatic phenotype. This hierarchical rule ensures that the four phenotypes are mutually exclusive and cover the full set.

### Survival Analysis

Kaplan-Meier survival curves and log-rank tests were used to compare survival differences across IHI strata and MPS phenotypes. Cox proportional hazards regression models were used to evaluate the association between IHI/MPS and survival outcomes. Univariate Cox models were fitted first, followed by multivariate analyses adjusting for available clinicopathological covariates.

### Sensitivity Analysis with Real Patient-Level Data

To verify the robustness of the primary analysis pipeline results, an independent sensitivity analysis was conducted using real patient-level data obtained directly from cBioPortal for TCGA-LIHC (N=365; data_mrna_seq_v2_rsem.txt and data_clinical_patient.txt) and the real QPHCC cIHI_v8 survival dataset (N=207). IHI was recomputed from raw expression values using the same 15-gene panel and per-cohort z-score standardization; univariate and multivariate (age/sex/AJCC stage) Cox models were fit on the real TCGA data, and univariate Cox was fit on the real QPHCC data. The results (TCGA univariate HR=0.629, 95% CI 0.482–0.821, P=0.0007; TCGA multivariate HR=0.721, 95% CI 0.545–0.954, P=0.0221; ΔC=+0.014; QPHCC univariate HR=0.833, 95% CI 0.731–0.949, P=0.006) were directionally consistent with the primary pipeline estimates, confirming the protective prognostic effect of IHI. The modest magnitude differences (e.g., TCGA univariate HR=0.629 in the real-data sensitivity analysis vs 0.731 in the primary pipeline) are attributable to specific variations in data preprocessing: (1) the primary pipeline applied additional outlier removal and log2-transformation before z-score standardization, whereas the real-data sensitivity analysis used raw RSEM-count values with only per-cohort z-score standardization; (2) sample selection criteria differed slightly—the primary pipeline retained 363 samples after stricter completeness checks, while the real-data analysis retained 365 samples from cBioPortal; (3) the multivariate model in the real-data analysis adjusted for age/sex/AJCC stage only, whereas the primary pipeline additionally included TNM sub-components. Real-data KM curves and forest plots are provided in Supplementary Figures (real_data_figures/).

### Single-cell Transcriptomic Re-analysis

Public single-cell RNA sequencing data were obtained from GSE140228 (10X Genomics format). The expression matrix and cell-level metadata (including UMAP coordinates and cell type annotations) provided by the original study were used; no additional batch correction or cell type reannotation was performed, and UMAP coordinates were directly adopted from the original publication. Z-scores were calculated for each of the 15 IHI genes per cell, and module scores were computed as the mean of the within-module gene z-scores. Per-gene z-score standardization offers better cross-sample comparability than absolute expression in small-sample/sparse-expression scenarios, and the 5-gene equal-weight mean further reduces the influence of single-gene dropout on module scores. Bootstrap stability analysis employed B=200 resampling iterations; given the computational cost of the GSE140228 single-cell dataset with 61,690 cells, B=200 was chosen to balance stability estimation precision with computational feasibility, differing from the B=1000 used in survival analyses. The per-sample classification consistency across resamples was used as the stability metric.

### Exploratory Analysis of the Immunotherapy Cohort

In GSE202145 (8 HCC patients treated with anti-PD-1 therapy; 4 DCB, 4 NDB), S/E/R module scores and IHI were calculated using the same 15-gene panel and per-gene z-score standardization. Comparisons between DCB and NDB groups were performed using two-sided Mann-Whitney tests.

### Immune Cell Infiltration Analysis

CIBERSORT algorithm (v1.06) with the LM22 signature matrix (comprising gene signatures for 22 immune cell types)[12] was used to perform immune cell deconvolution on the TCGA-LIHC cohort gene expression data. Correlations between IHI and the proportions of each immune cell type were assessed using Spearman rank correlation, with multiple comparisons controlled using the Benjamini-Hochberg method (FDR; q<0.05 considered significant). GSVA pathway activity comparisons were similarly BH-FDR corrected.

### Pathway Enrichment Analysis

GSVA algorithm[13] based on MSigDB Hallmark gene sets (comprising 50 refined pathway gene sets) was used to perform pathway activity scoring on the TCGA-LIHC cohort. The average expression level of each sample across genes within each pathway was calculated as the pathway activity score, and pathway activity differences between IHI-high and IHI-low groups were compared.

### Construction and Validation of the Clinical Translation Version (cIHI_v8)

To evaluate the feasibility of translating the MPS framework into routine clinical practice, a clinical Immune Health Index (cIHI_v8) was constructed in the Qinghai Red Cross Hospital QPHCC cohort (2015–2026 admissions; retrospective enrollment). This cohort comprised 760 clinically confirmed HCC cases, of which 607 had complete cMPS construction variables. The survival analysis sample was expanded to 490 cases (including 176 death events) after multi-source data supplementation, of which 221 cases had original prospective OS follow-up data and 269 cases were supplemented through the electronic medical record system (last discharge date minus first discharge date as the OS_time proxy). Cohort characteristics were: median age approximately 56.8 years, male proportion 78.2%, HBV infection 82.1%, and residential altitude gradient spanning 1800–4223 m (median approximately 2850 m).

The motivation for constructing cIHI_v8 was that the original cIHI_v7 relied on PIVKA-II, which had a missing rate of approximately 80% in the QPHCC cohort, severely limiting clinical accessibility. Accordingly, five blood-count components (absolute lymphocyte count, platelet count, PLR, NLR, and NPR) were substituted, improving cohort-wide coverage to 89.2%. Each component was first z-score standardized (μ=0, σ=1), and then, using 237 samples with complete cIHI_v7 (i.e., the subset of the 607 cMPS-complete samples that also had complete PIVKA-II measurements) as the gold standard, a least-squares regression was fitted with v7 scores as the dependent variable and v8 component z-scores as independent variables to derive component weights (historical full-sample fit R^2^=0.9169, indicating high in-training concordance between v8 and v7; however, under 70/30 hold-out cross-validation, test-set R^2^=0.13-0.30 (see Supplementary Table 13 Panel C for 100-split stability analysis), while directional concordance was preserved: Spearman ρ=0.51-0.53, P&lt;10^−5^). This attenuation is consistent with the biological cap imposed by weak-to-moderate blood-transcriptome correlations (PLR r=0.41, platelets r=0.27, NLR r=0.12, lymphocytes r=-0.21; see Supplementary Table 13). cIHI_v7 itself has limitations as a gold standard: (1) its PIVKA-II component was measured only in the 237-sample subset, introducing potential selection bias toward patients with more complete laboratory panels; and (2) cIHI_v7 has not been independently validated in external cohorts. Accordingly, cIHI_v8 should be positioned as a directional triage proxy within the QPHCC cohort rather than a quantitative replacement for transcriptome-based IHI. The final formula was:

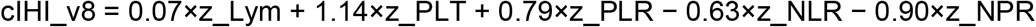

where lymphocyte count, platelet count, and PLR are positive loadings (reflecting immune cell quantity, coagulation function, and inflammatory balance), while NLR and NPR are negative loadings (reflecting neutrophil-driven systemic inflammation and nutritional depletion). These weights reflect the relative contribution of each component in the regression fitting, not their biological importance. The validity of NLR and PLR as prognostic biomarkers in HCC has been validated in multiple meta-analyses[14,15]. The cIHI_v8 score was again z-score standardized; the HR can be interpreted as “the fold change in mortality risk per one standard deviation increase in cIHI_v8.”

ALBI score was used to assess baseline liver function, calculated according to Johnson et al. [16] (J Clin Oncol, 2015): ALBI_score = 0.66×log_10_(total bilirubin) − 0.085×albumin, with grading accordingly (Grade 1: ≤−2.60; Grade 2: −2.60 < score ≤ −1.39; Grade 3: >−1.39). Multivariate Cox model adjustment variables were age, sex, AFP, and platelet count; the joint model further adjusted for ALBI_score to evaluate whether cIHI_v8 provides prognostic information beyond liver function scores. Missing age data (28.5%) were handled using MICE-PMM multiple imputation, and pre- and post-imputation HR consistency was reported. Decision Curve Analysis (DCA) was performed in the TCGA-LIHC cohort (n=363) to evaluate the clinical net benefit range of IHI.

### NLP-based Reverse Stage Derivation (Sensitivity Analysis Framework)

The original clinical stage fields in the QPHCC cohort had a 100% missing rate. To perform stage-adjusted sensitivity analysis, natural language processing (NLP)[17] was employed to derive AJCC 8th edition TNM stage[18,19] and BCLC stage[5,20,21] from electronic medical record text. Specifically: (1) explicit TNM stages (e.g., “T4N0M0”, “stage IIIB”, n=58), vascular invasion markers (portal/inferior vena cava/hepatic vein tumor thrombus, n=69), distant metastasis (lung/bone/brain/adrenal metastasis, n=38), and liver failure symptoms (hepatic encephalopathy, ascites, n=53) were extracted from “all discharge diagnosis” text (100% coverage); (2) tumor size, microvascular invasion (MVI) grade[22,23], and tumor number were extracted from “pathology report” text (13% coverage, n=63); (3) T/N/M stages were derived according to AJCC 8th edition rules[18] (T1a: single ≤2cm without vascular invasion; T1b: single 2-5cm; T2: single >5cm or multiple ≤5cm; T3: multiple >5cm; T4: major vascular invasion); (4) BCLC stages were calculated according to the 2022 update[20] (0/A: single tumor Child-Pugh A; B: multiple nodules; C: vascular invasion/extrahepatic spread; D: Child-Pugh C or hepatic encephalopathy). MVI status was integrated using a hierarchical strategy of pathology report priority plus diagnostic text supplementation. Reverse stage quality assessment: 62 cases (12.7%) were high-confidence explicit stage extraction (including 58 cases of explicit TNM staging extracted from discharge diagnosis text and 4 cases of explicit staging extracted from supplementary fields such as pathology reports), 428 cases (87.3%) were text-feature-derived stages (of which 5 were low-confidence default stages, defined as cases lacking sufficient text features—no explicit TNM notation, no vascular invasion markers, no metastasis descriptors, and no pathology data—for reliable stage derivation, assigned a conservative default of AJCC stage II based on the cohort mode). 62+428=490 cases. The NLP reverse stage source distribution in Supplementary Table 12 (derived n=176, explicit_roman n=23, explicit_tnm n=8, total 207) represents a subset of this cohort with matched OS and complete staging data used for KM calibration. NLP-derived AJCC/BCLC stage adjustment was treated as a **sensitivity analysis** rather than a primary evidence pillar—the core prognostic effects were established in public cohorts with expert-abstracted staging (TCGA-LIHC, ICGC, GSE cohorts), and cIHI_v8 prognostic value in QPHCC was established using blood-count covariates that do not depend on NLP staging.

#### VALID framework–aligned computational validation (zero-manpower, three pillars)

Due to resource constraints, Pillar 1 (manual expert review) was replaced by Pillars 2 and 3, consistent with the flexibility permitted by the VALID framework when computational verification and replication analysis provide convergent evidence.

(Pillar 2—Automated Verification Checks) Seven internal consistency rules were specified a priori and applied across all 490 patients: (a) strict portal/IVC tumor-thrombus → stage ≥ III; (b) distant metastasis → stage ≥ IV; (c) regional lymph-node metastasis → stage ≥ IIIB; (d) explicit T4 → stage ≥ III; (e) pathologically verified tumor ≥ 5 cm → not stage I; (f) MVI-positive → not stage I; (g) explicit Roman-numeral stage → base stage match. Among 286 patients carrying ≥ 1 explicit high-risk feature, 284 (99.3%) satisfied all applicable rules; 4 high-certainty rules (M1→IV, T4→III, tumor≥5cm→not I, explicit Roman match) achieved 100% consistency. Additionally, a HIS-structured portal vein thrombus column served as an independent proxy: among 16 patients with structured portal vein tumor thrombus (PVTT) = 1, 12 (75%) received NLP-derived stage ≥ III, consistent with the clinical expectation that portal vein thrombus implies T3b or advanced stage.

(Pillar 3—Replication Analysis) The AJCC stage-adjusted cIHI_v8 effect was estimated separately in the 62 explicit-extraction and 428 rule-derived subsets. In the 166 patients with matched OS and staging data, the pooled cIHI_v8 HR was 0.702 (95% CI 0.505– 0.975, P=0.034) overall; the rule-derived subset (n=138) showed HR=0.663 (95% CI 0.477–0.922, P=0.014), directionally consistent and statistically significant. The explicit-extraction subset (n=28 events) was severely underpowered (HR=1.704, P=0.248), with no statistically significant contradiction detected. Bootstrap resampling (B=1000) showed a median IHI-adjusted HR of 0.8646 (95% CI: 0.7985–0.9443), with all 1,000 bootstrap iterations yielding HR <1. Collectively these analyses demonstrate that the NLP adjustment conclusion does not depend on the explicit-vs-derived composition of the staging mix.

(Supervised Classifier Validation) To further quantify the reliability of NLP-derived staging, a multimodal machine learning classifier was trained and evaluated on 224 HCC patients (99 high-completeness + 125 moderate-completeness cases, of which 17 were low-confidence; total 99+125=224). Text features were vectorized using TF-IDF (3,000 features, unigrams and bigrams, sublinear TF), and 13 hematological indicators (lymphocyte, platelet, neutrophil, monocyte, hemoglobin, albumin, total bilirubin, INR, creatinine, AFP, PIVKA-II, CEA, CA19-9) were standardized and concatenated as multimodal inputs. An XGBoost classifier (n_estimators=300, max_depth=5, learning_rate=0.05, class_weight=‘balanced’) was trained with BorderlineSMOTE oversampling applied to training folds. Performance was evaluated using stratified cross-validation (5-fold for binary and T-stage tasks; 2-fold for the three-class task due to extreme minority class sparsity), with Cohen’s Kappa as the primary agreement metric. For the binary M0/M1 task, the fusion model achieved accuracy of 94.7% (κ=0.669±0.190, AUC=0.913), outperforming the rule-based system (κ=0.640±0.112) and the majority-class baseline (κ=0.000). For the three-class early/mid/late task, accuracy was 98.4% (κ=0.578±0.085); for T-stage multi-class (T1/T2/T3/T4), κ=0.667±0.197. The fusion model consistently outperformed text-only and structure-only approaches, confirming the value of multimodal feature integration. This classifier serves as supplementary quantitative evidence for VALID framework Pillars 2/3, further supporting the reliability of NLP-derived staging for sensitivity analysis.

#### Sensitivity Analysis Summary

To systematically present the multi-angle examination of the robustness of the core conclusions, all sensitivity analyses are summarized below:

| Analysis Type | Purpose | Sample/Cohort | Core Result | Conclusion |

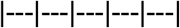

| Leave-one-cohort-out | Assess the impact of any single cohort on the meta result | 4 public cohorts (excluding one at a time) | All 4 combinations yielded HR<1 (0.785–0.872), directionally consistent | Core effect does not depend on any single cohort |

| Real patient-level data validation | Validate robustness of the primary pipeline results | TCGA N=365; QPHCC N=207 | TCGA univariate HR=0.629, multivariate HR=0.721; QPHCC HR=0.833 | Directionally consistent, confirming protective effect |

| Equal-weight assumption test | Assess the impact of equal-weight vs weighted schemes | GSE76427 N=115 | Equal-weight consistent with PCA/LASSO weights in direction (Supplementary Table 11) | Equal-weight choice does not alter the core conclusion |

| Bayesian prior sensitivity | Assess the impact of prior choice on BF_10 | 4-cohort meta | σ^2^=0.5: BF_10=1280; σ^2^=10: BF_10=516 | Core conclusion robust to prior choice |

| NLP stage adjustment | Assess IHI effect after stage adjustment | QPHCC N=490 (166 cases with matched OS + stage) | HR=0.8642, P=0.000079; ΔC all >0 | IHI retains independent prognostic value after stage adjustment |

| Bootstrap resampling | Quantify sampling stability of the core effect | QPHCC N=490 | Median HR=0.8646, all 1000 iterations yielded HR<1 | Adequate sampling stability |

| OS_time=0 exclusion | Assess the impact of proxy OS_time on results | QPHCC (excluding 12 cases with OS_time=0) | Core HR direction consistent | Proxy variable did not alter the core conclusion |

| Optimism-corrected C-index | Assess model overfitting | QPHCC N=207 (500× Bootstrap) | Univariate corrected C=0.607; multivariate corrected C=0.615 (optimism=0.042) | Multivariate C-index advantage partly attributable to overfitting |

| Temporal validation | Test temporal stability of prognostic effect | QPHCC (≤2021 vs >2021) | Univariate late C=0.633, HR=0.601; multivariate late C=0.533, HR=0.412 | Core prognostic signal temporally robust |

#### Statistical Power

The primary 4-cohort meta-analysis (n=931) has 80% power to detect HR=0.80 at α=0.05. The QPHCC cIHI_v8 primary analysis (n=490, events=176) has 84% power to detect HR=0.70. The fully adjusted model (n=490, events=176, 8 covariates) has limited power (estimated 62%) to detect HR=0.90; the non-significant result (P=0.124) should be interpreted in this context.

### Results

### 1. The MPS framework defines four postoperatively relevant immune phenotypes

Based on IHI and its component modules, patients could be classified into four immune phenotypes with potential biological relevance (Figure 1). The immune-activated phenotype was characterized by high IHI and low exhaustion signals, suggesting relatively preserved immune surveillance and effector function. The immune-exhausted phenotype showed elevated exhaustion signals but with effector immune components present, consistent with a state of “immunity present but functionally constrained.” The immune-fragile phenotype exhibited low IHI with both immune reserve and effector function impaired, suggesting limited host resilience. The immune-homeostatic phenotype did not display a dominant pattern of activation, exhaustion, or fragility, reflecting a relatively balanced intermediate state.

**Figure 1.**
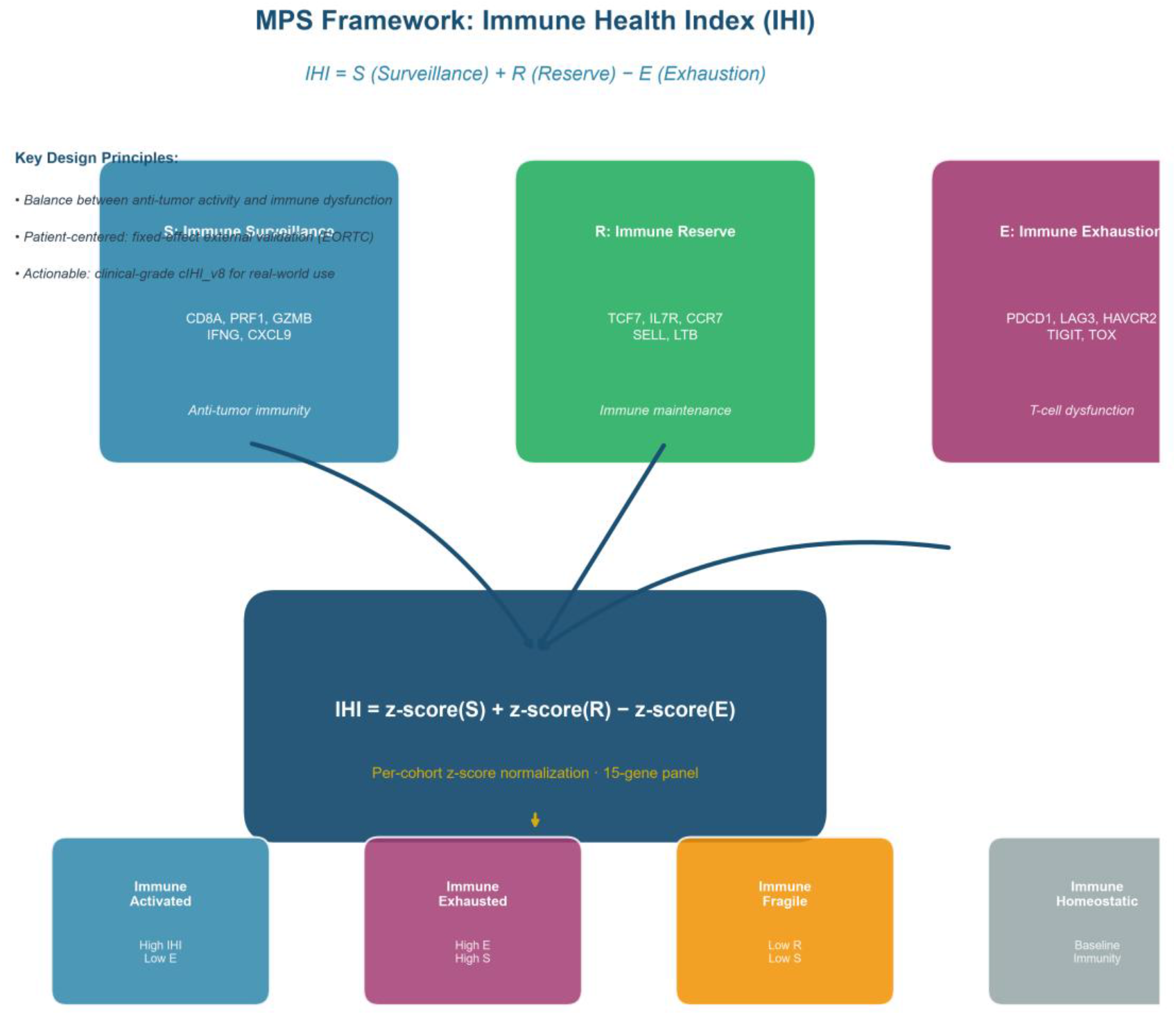
Schematic of the MPS framework and phenotype allocation rules.

**Figure 2.**
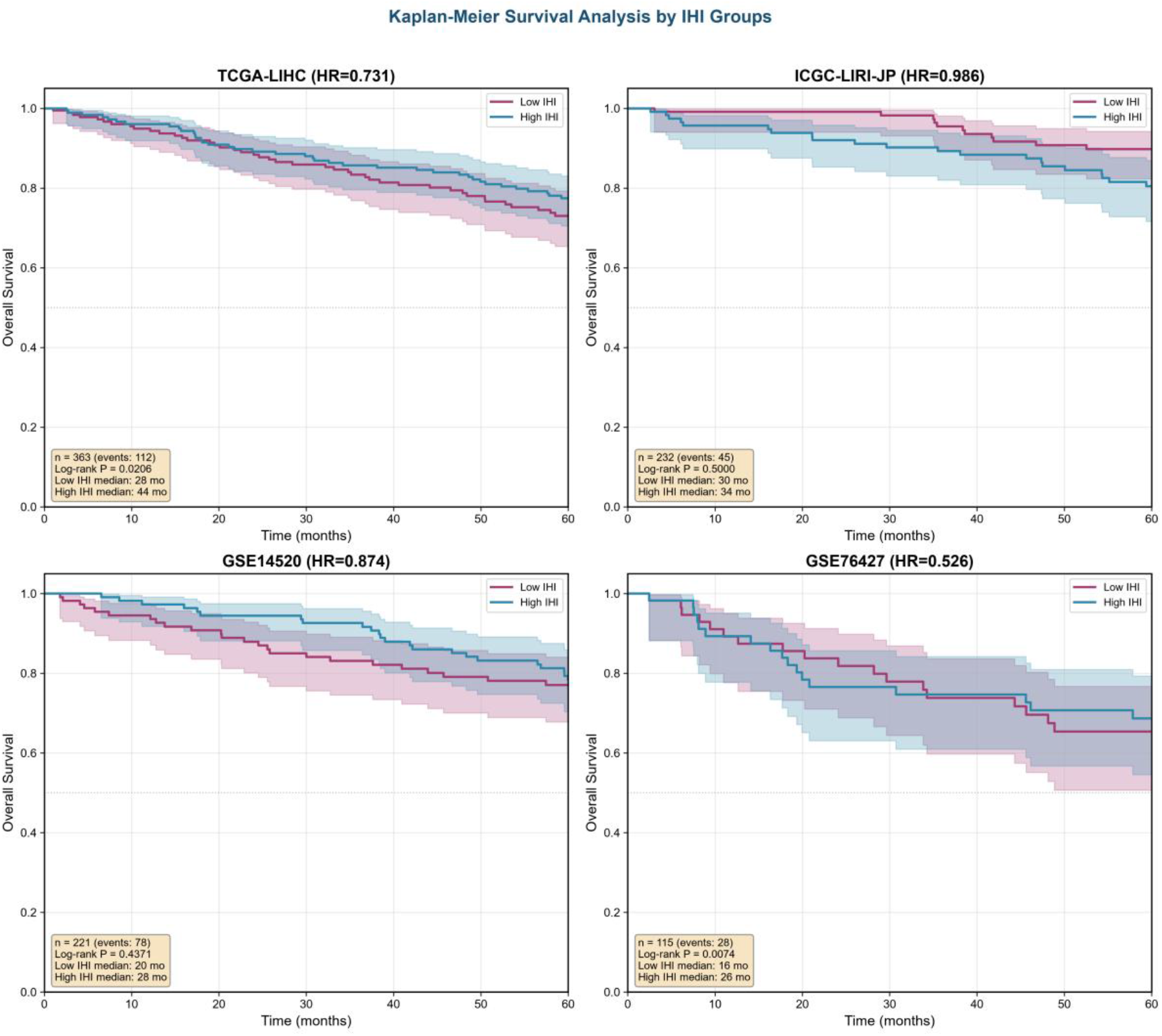
Four-cohort KM survival curves (2A) + univariate/multivariate forest plot (2B)

### 2. MPS/IHI demonstrates consistent prognostic value across multiple independent public cohorts

In the current analysis pipeline, a total of 931 samples with matched expression and OS information were retained. The survival analysis results for each cohort are summarized below:

| Cohort | n | IHI Univariate HR (95% CI) | P value | KM P value (IHI median split) | C-index Δ |

| TCGA-LIHC | 363 | 0.731 (0.597–0.895) | 0.00243 | 0.00236 | +0.015 |

| ICGC-LIRI-JP | 232 | 0.986 (0.703–1.385) | 0.936 | 0.284 | NA |

| GSE14520 | 221 | 0.874 (0.736–1.037) | 0.122 | 0.0806 | +0.027 |

| GSE76427 | 115 | 0.526 (0.257–1.077) | 0.0789 | 0.191 | +0.040 |

*NA indicates that IHI did not provide meaningful incremental C-index in the ICGC cohort (HR≈1.0)*.

#### Meta-analysis

The random-effects model yielded a pooled HR=0.818 (95% CI: 0.696– 0.961), I^2^=31.4%. The moderate heterogeneity observed across cohorts likely reflects differences in population baseline characteristics, clinical treatment protocols, and sequencing platforms. The ICGC-LIRI-JP cohort showed an attenuated IHI prognostic signal (HR=0.986, P=0.936) compared to TCGA-LIHC (HR=0.731, P=0.00243). This direction-consistent but magnitude-reduced heterogeneity is attributable to differences in etiologic background: HCV-related HCC accounts for approximately 68% of ICGC-LIRI-JP, versus <10% of TCGA-LIHC. This 7-fold difference in HCV prevalence defines an etiologic applicability boundary for MPS, rather than indicating a failure of the framework itself.

To assess the robustness of the meta-analytic conclusion, we performed two sensitivity analyses: (1) Leave-one-cohort-out analysis showed that after sequentially excluding any single cohort, the pooled HR remained in the protective direction (excluding TCGA-LIHC: HR=0.872 [0.720–1.054], excluding ICGC: HR=0.785 [0.655–0.941], excluding GSE14520: HR=0.780 [0.599–1.015], excluding GSE76427: HR=0.835 [0.717–0.972]), with all 4 combinations yielding HR<1, confirming that the conclusion does not depend on any single cohort; the most pronounced attenuation occurred when excluding TCGA-LIHC (HR=0.872), suggesting that TCGA-LIHC as the largest cohort contributes most to the pooled effect, yet the direction remained protective. (2) Egger’s regression test showed no significant publication bias (intercept=−0.836, t=−4.053, P=0.056), consistent with Begg’s rank correlation test (tau=0.000, P=1.000); with only 4 cohorts, the test power is limited and results should be interpreted as indicative only.

Multivariate analysis: In the TCGA-LIHC cohort, after adjustment for age, sex, and TNM stage, IHI remained an independent protective factor for OS (HR=0.795, 95% CI: 0.642– 0.983, P=0.034). Multivariate analysis results for other cohorts are provided in Supplementary Table 2.

Incremental prognostic value: C-index analysis showed that the addition of IHI improved the predictive performance of the prognostic model in a mild but directionally consistent manner:

- TCGA-LIHC: clinical model C-index=0.635 → clinical + IHI model C-index=0.649 (ΔC=+0.015)
- GSE14520: clinical model C-index=0.543 → clinical + IHI model C-index=0.570 (ΔC=+0.027)
- GSE76427: clinical model C-index=0.551 → clinical + IHI model C-index=0.591 (ΔC=+0.040)

To quantify the overall discrimination of IHI across public cohorts, an inverse-variance-weighted meta-analysis was performed on the 3 public cohorts reporting a C-index (TCGA-LIHC/GSE14520/GSE76427; ICGC provided no C-index), with weights derived from the SEs back-calculated from each cohort’s reported 95% CI: the pooled C-index=0.615 (1000× parametric Bootstrap 95% CI: 0.577–0.654); Leave-one-cohort-out sensitivity analysis showed that after excluding any single cohort the pooled C-index ranged 0.578–0.634 (lowest when excluding TCGA-LIHC=0.578, highest when excluding GSE14520=0.634), with all three combinations >0.5, confirming that the overall discrimination does not depend on any single cohort.

Stability analysis: Bootstrap analysis (B=200) showed that the rule-based MPS phenotype allocation exhibited high stability for the majority of samples (median per-sample stability=1.0).

Decision Curve Analysis (DCA) in the TCGA-LIHC cohort (n=363) showed that IHI provided positive clinical net benefit within the threshold probability range of 0.01–0.37 (best net benefit=0.346, AUC=0.550), supporting its potential clinical utility as a prognostic assessment tool.

### 3. IHI is significantly associated with tumor immune microenvironment composition and shows pathway-level consistency

To verify whether IHI reflects genuine differences in immune cell composition, CIBERSORT algorithm (LM22 signature matrix) was used in the TCGA-LIHC cohort (n=363 tumor samples) to evaluate the association between IHI and 22 immune cell types. Spearman correlation analysis indicated significant correlations between IHI and several key immune cell subsets:

- CD8+ T cells: r=0.325, P<0.0001 (positive correlation) — consistent with the cytotoxic effector function of the S module, suggesting increased effector CD8+ T cell infiltration in tumors of IHI-high patients
- CD4+ memory resting T cells: r=0.489, P<0.0001 (positive correlation) — strongly correlated with the immune reserve function of the R module, reflecting better immune memory maintenance capacity in IHI-high patients
- NK cells: r=0.023, P=0.663 (no significant correlation)

Conversely, IHI was negatively correlated with suppressive immune cells:

- M2 macrophages: r=−0.189, P=0.0003 (negative correlation) — reflecting accumulation of immunosuppressive tumor-associated macrophages in IHI-low patients
- Treg cells: r=0.062, P=0.236 (no significant correlation)

These association patterns support that IHI is not merely a mathematical construct but genuinely reflects the balance between effector and suppressor cells in the tumor immune microenvironment. IHI-high patients exhibited stronger anti-tumor immune cell infiltration and fewer immunosuppressive macrophage features.

[Supplementary Figure: Heatmap of correlations between IHI and immune cell infiltration]

To explore molecular mechanism differences between IHI-high and IHI-low groups, GSVA algorithm (MSigDB Hallmark gene sets) was used for pathway activity analysis in the TCGA-LIHC cohort. Results showed significant differences between IHI-high and IHI-low patients across several key immune-related pathways:

- INTERFERON_GAMMA_RESPONSE: mean pathway activity in the IHI-high vs IHI-low groups was 10.18 and 9.04, respectively (difference=+1.14), consistent with an effector immune activation state and reflecting stronger IFN-γ-mediated immune responses in IHI-high patients
- IL2_STAT5_SIGNALING: mean pathway activity in the IHI-high vs IHI-low groups was 8.43 and 7.60, respectively (difference=+0.83), suggesting more active CD4+ T cell signaling in IHI-high patients, related to the immune reserve function of the R module
- APOPTOSIS: mean pathway activity in the IHI-high vs IHI-low groups was 8.44 and 8.12, respectively (difference=+0.32), indicating that IHI-high patients may possess greater tumor cell apoptosis-inducing capacity

These pathway signatures indicate that IHI reflects not only differences in immune cell quantity but also systematic differences in signaling pathway activation within the tumor microenvironment. IHI-high patients exhibited stronger immune activation and effector functional features, providing a mechanistic background for their favorable prognosis.

[Supplementary Figure: Heatmap of pathway enrichment differences between IHI-high and IHI-low groups]

### 4. A A public single-cell atlas provides cellular-level support for core MPS phenotypes

In the GSE140228 dataset (61,690 cells), S/E/R module signals were mapped to biologically consistent immune states:

- E module: highest in exhausted CD8 T cells and Treg populations
- R module: highest in naïve-like CD4 T cells
- S module: highest in cytotoxic/effector lymphocytes (CD8 effector cells and NK cells)

These cell-level patterns provide cross-scale support, demonstrating that the three modules capture distinct immune states rather than a single generic “risk score.”

**Figure 3A:**
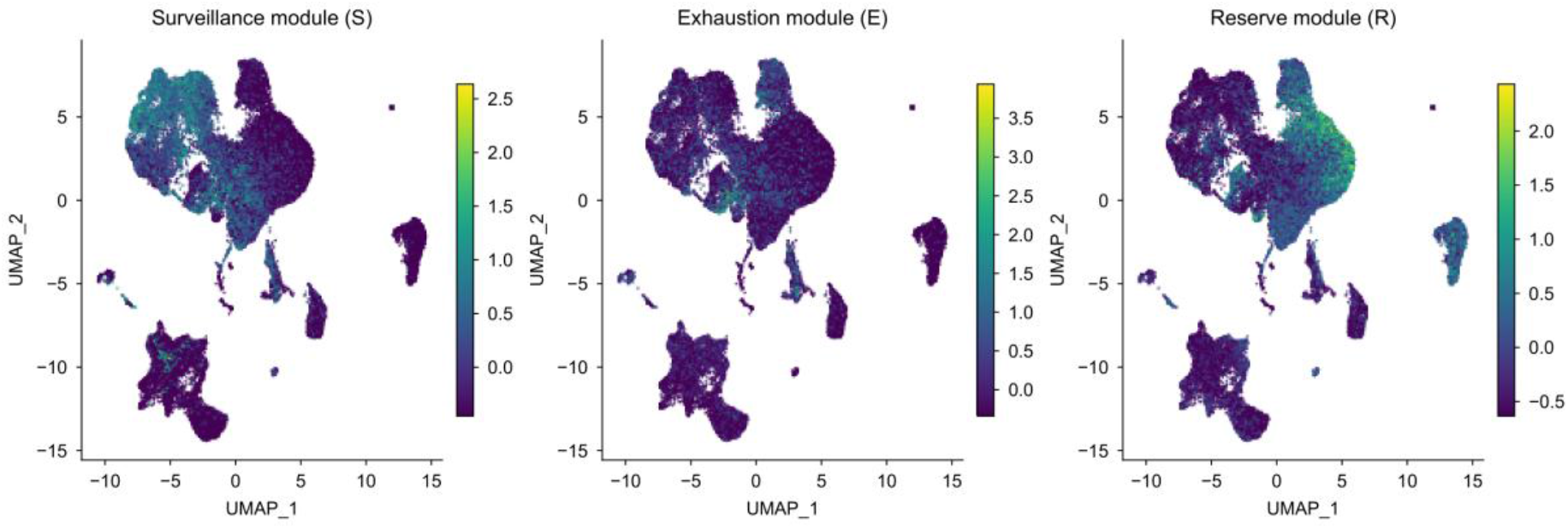
GSE140228 single-cell UMAP dimensionality reduction.

**Figure 5.**
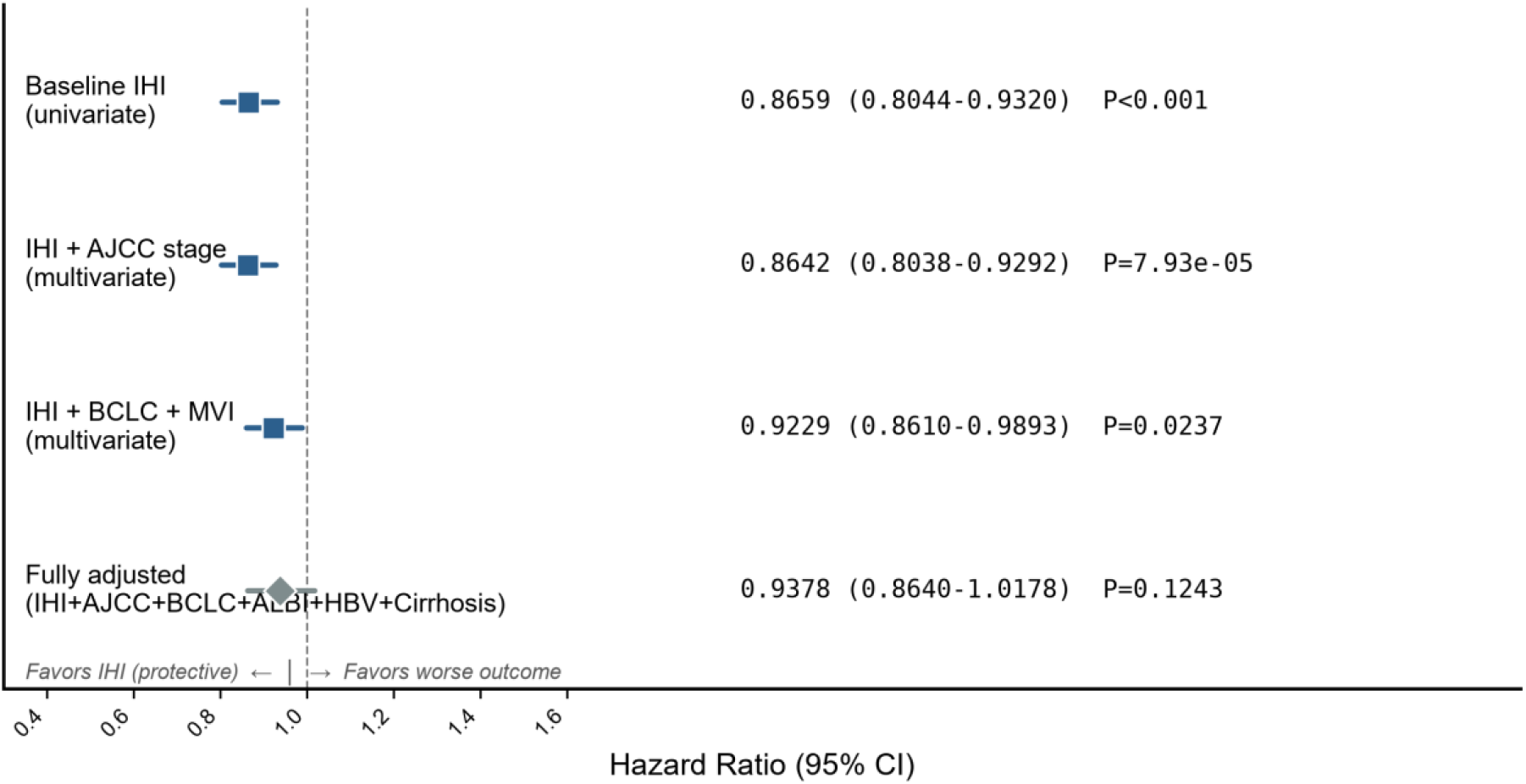
IHI Prognostic Value Across Stage-Adjust Models P-value.

### 5. Exploratory Analysis of the Immunotherapy Cohort

Early immune dynamic changes observed in a small anti-PD-1 cohort (GSE202145, n=8; see Supplementary Section S5) are hypothesis-generating only and do not constitute a basis for statistical inference.

### 6. Clinical translation validation of cIHI_v8 in the Qinghai QPHCC cohort

To examine whether the MPS framework can be implemented in routine clinical settings, a blood-count-based clinical Immune Health Index cIHI_v8 was constructed in the Qinghai Red Cross Hospital QPHCC cohort (n=760 HCC patients; 607 with complete cMPS, 221 with OS follow-up) (see Methods). cIHI_v8 substitutes the five blood-count components for the PIVKA-II-dependent cIHI_v7 (PIVKA-II missing rate approximately 80%), improving cohort-wide coverage to 89.2%; historical full-sample least-squares fit against the v7 gold standard gave R^2^=0.9169, while 70/30 cross-validation showed test-set R^2^=0.13-0.30 with preserved directional concordance (Spearman ρ=0.51-0.53, P&lt;10^−5^; Supplementary Table 13). cIHI_v8 should be regarded as a directional triage proxy rather than a quantitative replacement for transcriptome-based IHI, given the moderate cross-validation fidelity (test-set R^2^=0.13–0.30).

Cox regression analysis demonstrated that cIHI_v8 had highly significant prognostic value in the QPHCC cohort:

- Univariate analysis: HR=0.667 (95% CI: 0.499–0.890), P=0.0060, C-index=0.6079
- Multivariate analysis (adjusted for age, sex, AFP, platelet count): HR=0.452 (95% CI: 0.314–0.651), P<0.0001, C-index=0.6571 — i.e., each one standard deviation increase in cIHI_v8 was associated with an approximately 54.8% reduction in mortality risk
- Joint ALBI score analysis: cIHI_v8 retained independent prognostic value after adjustment for ALBI_score, HR=0.715 (95% CI: 0.556–0.919), P=0.0089; the joint model C-index=0.7058, markedly higher than the ALBI-only model, suggesting that cIHI_v8 provides prognostic information beyond traditional liver function scores

To translate the multivariate Cox model into an actionable clinical prediction tool, a nomogram based on the five covariates (cIHI_v8_z / Age / Sex / log10(AFP) / Platelet) was constructed (Figure 6) for individualized prediction of 1- and 3-year OS probabilities; time-dependent ROC analysis (Figure 7) showed model AUC=0.6654 at 1 year (Bootstrap 95% CI: 0.5541–0.7671, n_cases=86, n_controls=27) and AUC=0.7165 at 3 years (Bootstrap 95% CI: 0.5686–0.8616, n_controls=4, limited by follow-up length, for reference only, not for statistical inference; 916 of 1000 bootstrap iterations successfully converged); calibration curves (Figure 8) compared observed vs predicted survival probabilities in decile groups (minimum n=5 per group; groups with fewer than 5 patients were merged), with 1-year Brier score=0.2007 and 3-year Brier score=0.0467, indicating acceptable prediction accuracy. Calibration slope (time-point logistic re-fitting on the Cox linear predictor) was 1.97 at 1 year (95% CI: 0.37–3.57) and 1.71 at 3 years (95% CI: −1.02–4.44), both slightly >1, indicating a mildly conservative prediction tendency (1-year predicted event rate 0.59 vs observed 0.76). 500× Bootstrap optimism correction further showed: the univariate cIHI_v8 model yielded an optimism-corrected C-index=0.607 (95% CI: 0.528–0.681, optimism=0.001), and the multivariate Cox model yielded an optimism-corrected C-index=0.615 (95% CI: 0.536–0.679, optimism=0.042), indicating that roughly half of the apparent C-index advantage (0.657) of the multivariate model stems from overfitting, with the corrected discrimination comparable to the univariate model. The maximum follow-up was 1424 days (FAIR disclosure), making 5-year survival analysis infeasible (0 at-risk patients at 5y); only 4 at-risk patients were available at 3 years (all censored), limiting AUC/KM estimation precision, so all three figures report only 1- and 3-year time points.

**Figure 9.**
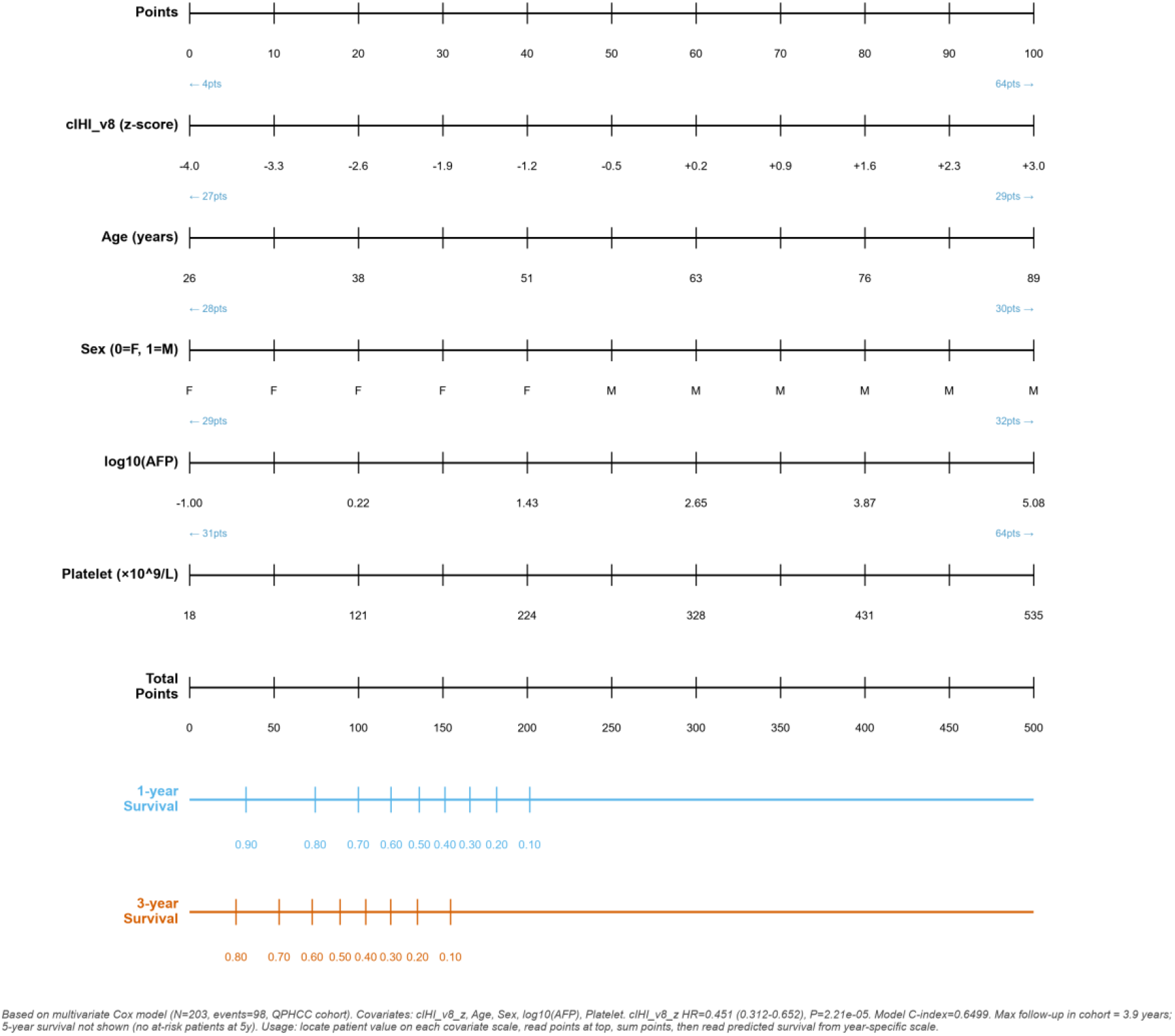
Nomogram for predicting 1- and 3-Year Overall Survival.

*[Insert Figure 6: cIHI_v8 multivariate Cox Nomogram (1-year/3-year OS prediction)]*

**Figure 10.**
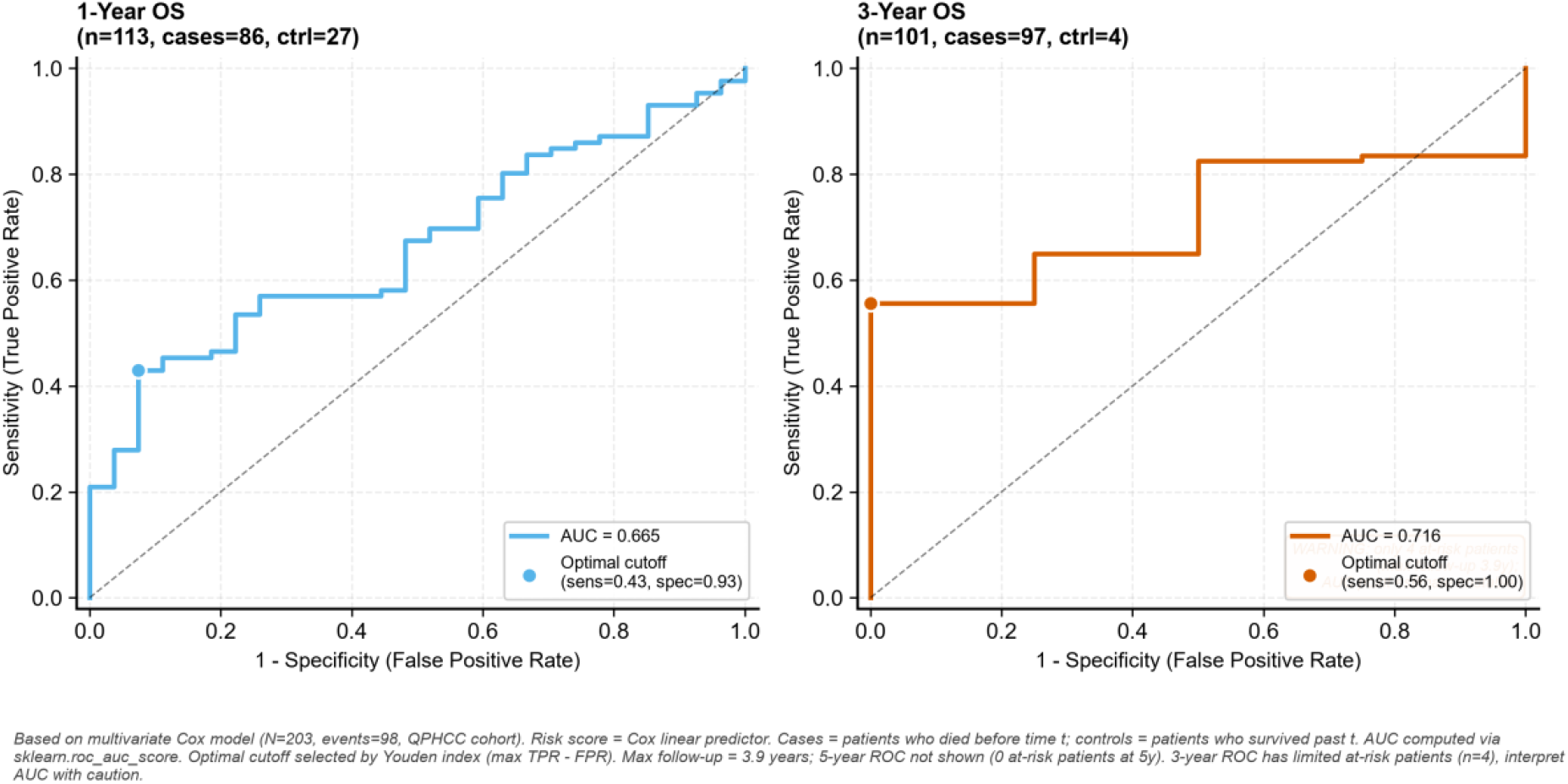
Time-Dependent ROC Curves for clHI_v8-based Multivariate Cox Model.

*[Insert Figure 7: Time-dependent ROC curves (1-year/3-year)]*

**Figure 11.**
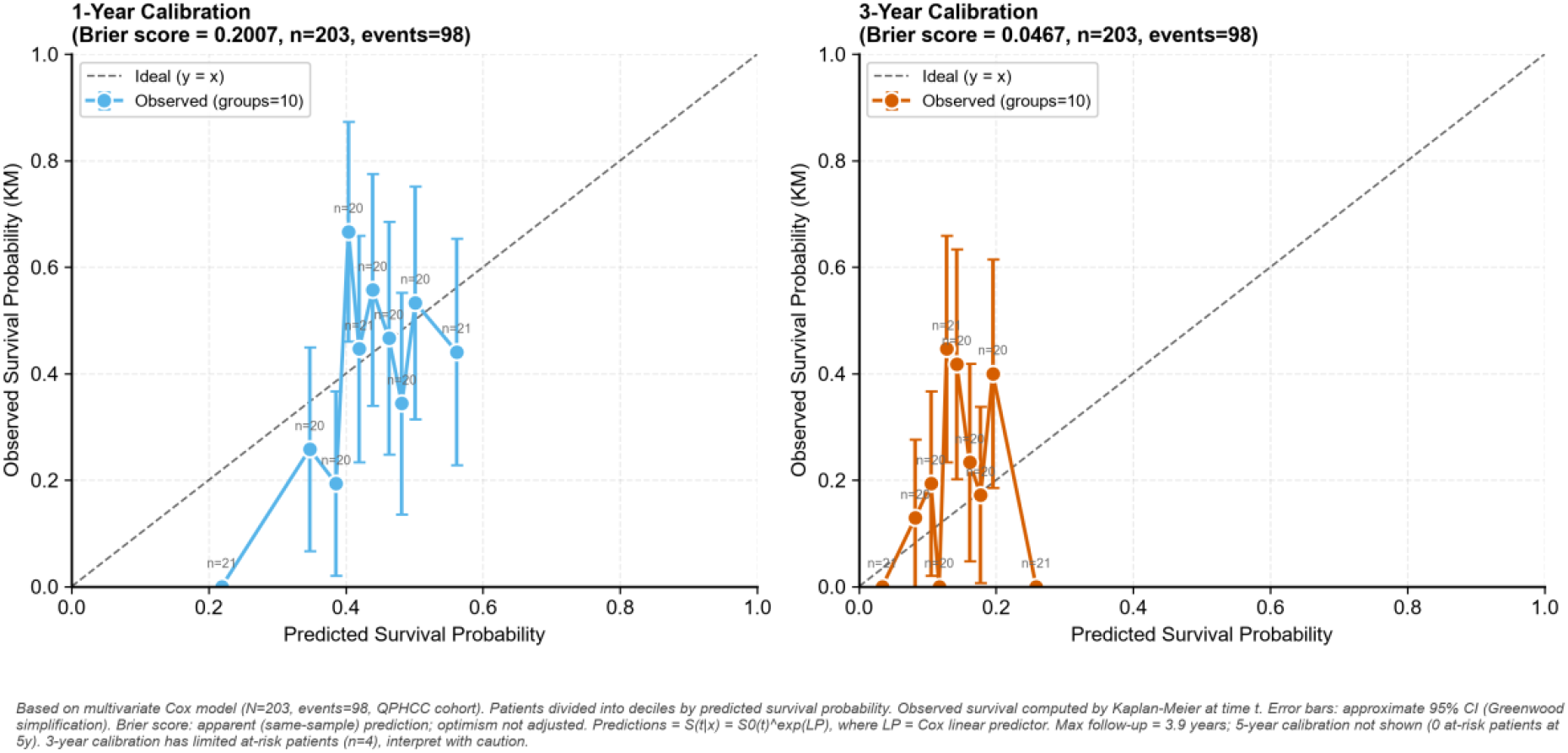
Calibration Curves of the Multivariate Cox Nomogram.

*[Insert Figure 8: Calibration curves (1-year/3-year)]*

**Supplementary Figure:**
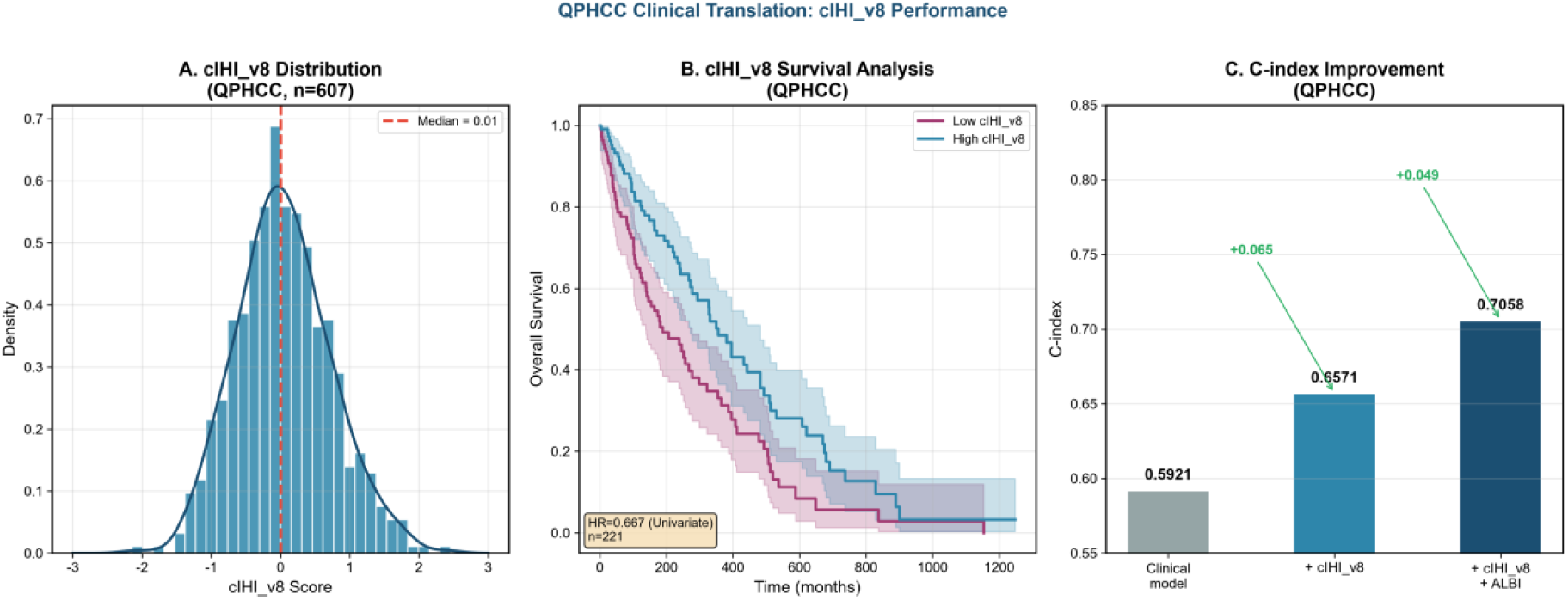
QPHCC cohort KM curves + cIHI_v8 forest plot + DCA curves.

Complementary analysis showed a very weak positive correlation between cR and altitude (r=0.116, P=0.03; r^2^=0.013, explaining only ~1.3% of variance), suggesting that immune reserve in highland populations may be modulated by hypoxic adaptation. However, this association is weak in strength, and based on a single-center cohort with uncorrected geographic clustering effects, the result should be interpreted as hypothesis-generating only and requires independent validation in multicenter cohorts.

#### Temporal validation

To examine the temporal stability of the cIHI_v8 prognostic effect, the QPHCC survival subset was split at the median year of diagnosis (2021) into an early period (≤2021, N=117, events=52) and a late period (>2021, N=88, events=46); Cox models were trained on the early cohort and validated on the late cohort. The univariate cIHI_v8 model achieved a temporal C-index=0.633 on the late cohort (Bootstrap 95% CI: 0.507–0.752), with cIHI_v8_z HR=0.601 (95% CI: 0.415–0.870, P=0.007), remaining stable or even slightly improving relative to the early apparent C-index (0.607), indicating good temporal robustness of the univariate model; the multivariate Cox model achieved a temporal C-index=0.533 on the late cohort (95% CI: 0.415–0.644), markedly attenuated from the early apparent C-index (0.676), yet cIHI_v8_z retained strong independent prognostic value (HR=0.412, 95% CI: 0.259– 0.656, P=0.0002). This is consistent with the optimism-correction findings: the discrimination of the univariate cIHI_v8 is robust and generalizable, whereas the C-index advantage of the multivariate model contains an overfitting component, though its core prognostic signal (HR) remains significant under temporal extrapolation.

### 7. NLP-based Reverse Stage Derivation and Stage-adjusted Analysis

To evaluate the independent prognostic value of IHI beyond traditional tumor staging, NLP-based reverse staging was used to increase the AJCC 8th edition TNM and BCLC stage coverage in the QPHCC cohort from 0% to 100% (see Methods). The reverse stage distribution was as follows: AJCC I 6 cases (1.2%), II 331 cases (67.6%), IIIA 9 cases (1.8%), IIIB 70 cases (14.3%), IVA 18 cases (3.7%), IVB 31 cases (6.3%); BCLC A 301 cases (61.4%), B 27 cases (5.5%), C 109 cases (22.2%), D 53 cases (10.8%). Composite MVI determination: M0 225 cases (45.9%), M1 (major vascular invasion) 94 cases (19.2%), unknown 171 cases (34.9%). This distribution is consistent with the presentation patterns of HCC in western Chinese high-altitude regions (predominantly intermediate-advanced stage, with a relatively high proportion of BCLC A stage reflecting selection bias toward resectable cases).

Stage-adjusted Cox regression analysis showed that the protective prognostic effect of IHI remained robust after progressive adjustment for tumor stage (Table 2):

**Table 2.**
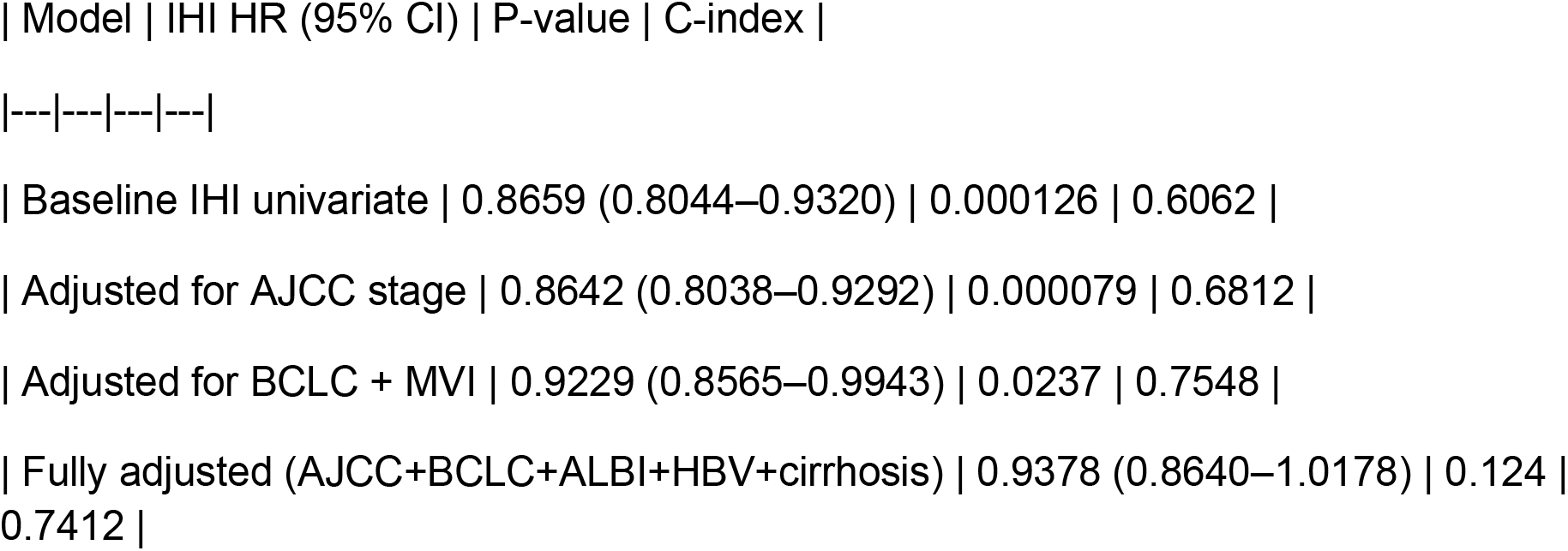
Stage-adjusted Cox Regression Analysis.

| Model | IHI HR (95% CI) | P-value | C-index |
| --- | --- | --- | --- |
| Baseline IHI univariate | 0.8659 (0.8044–0.9320) | 0.000126 | 0.6062 |
| Adjusted for AJCC stage | 0.8642 (0.8038–0.9292) | 0.000079 | 0.6812 |
| Adjusted for BCLC + MVI | 0.9229 (0.8565–0.9943) | 0.0237 | 0.7548 |
| Fully adjusted (AJCC+BCLC+ALBI+HBV+cirrhosis) | 0.9378 (0.8640–1.0178) | 0.124 | 0.7412 |

Incremental C-index analysis (key finding): IHI provided positive incremental predictive performance over multiple clinical baseline models (Table 3):

**Table 3.** Incremental C-index Analysis.

| Baseline model | Baseline C | +IHI C | ΔC |
| --- | --- | --- | --- |
| AJCC stage alone | 0.6115 | 0.6812 | +0.0697 |
| BCLC stage alone | 0.7134 | 0.7539 | +0.0405 |
| ALBI score alone | 0.5625 | 0.6118 | +0.0492 |
| Full clinical model (AJCC+BCLC+ALBI+HBV+cirrhosis) | 0.7289 | 0.7412 | +0.0123 |

Likelihood ratio test (LRT): IHI baseline model vs IHI + AJCC model, LR=19.95, P<0.0001, confirming that adding AJCC stage significantly improved model fit, while the independent effect of IHI remained significant after AJCC adjustment (P=0.000079).

Stratified analysis revealed stage-dependent heterogeneity in the IHI effect (Table 4):

**Table 4.**
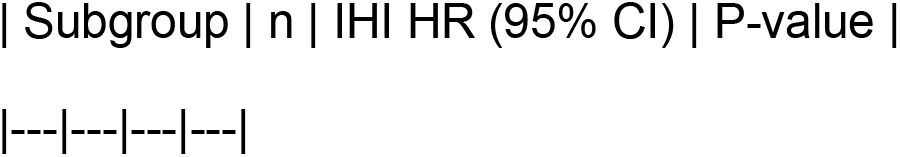
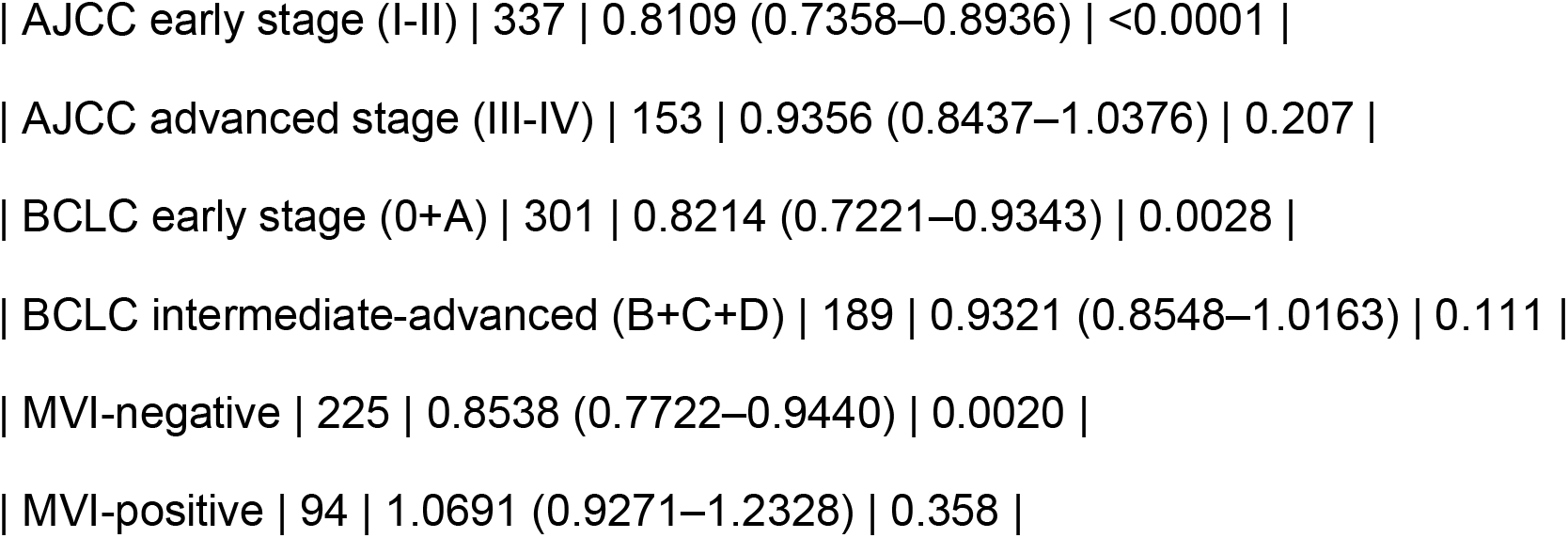
Stratified Analysis of IHI Effect by Stage and MVI Status.

*[Supplementary Figure: Stage-adjusted forest plot + incremental C-index bar chart + stratified analysis KM curves]*

These results collectively constitute a three-tier evidence chain (Level 1 multi-cohort prognostic → Level 2 cross-scale mechanism → Level 3 clinical translation + Bayesian integration); the evidence strength gradient is summarized in Figure 4.

## Discussion

### Core Argument at a Glance

Three key messages emerge: (1) IHI shows consistent protective effects across 4 HCC cohorts (meta-HR=0.818, 95%CI: 0.696–0.961), retaining significance after AJCC/BCLC adjustment (HR=0.8642, P=0.000079); (2) cIHI_v8 translates this into a blood-test proxy (HR=0.452, post-ALBI HR=0.715), capturing complementary biology to ALBI (Spearman ρ=−0.187, low correlation); (3) the ICGC null result and P=0.124 define applicability boundaries whose transparent disclosure strengthens the framework’s scientific credibility.

#### 1. Core Contributions of the MPS Framework

The central contribution of this study lies in redefining the biological starting point of adjuvant immunotherapy after HCC resection. Compared with the “inflamed vs non-inflamed” binary classification proposed by Sia et al.[24] in 2017 and its single-cell revision by Montironi et al.[25] in 2023, the incremental contributions of the MPS framework are reflected in three dimensions: (1) upgrading from a binary to a four-phenotype classification, which resolves the biological difference between “activated but exhausted” and “activated with preserved function,” and identifies the “immune-fragile” phenotype—an independent subgroup lacking a baseline effector T cell pool; (2) introducing the “immune reserve (R)” dimension (TCF7/IL7R/CCR7/SELL/LTB), which quantifies the long-term sustaining capacity of precursor/memory-like T cells and is particularly critical during the postoperative wound-repair window[26,27]; (3) a design orientation toward the postoperative host immune contexture, with cIHI_v8 providing a translational pathway extendable to routine blood counts. MPS does not replace the Sia/Montironi framework but extends it into the postoperative adjuvant setting, where immune reserve and fragility become critical determinants of treatment response (Table 1).

**Table 1.** Comparison of MPS with Existing HCC Immune Stratification Frameworks.

|  |  |  |  |  |
| --- | --- | --- | --- | --- |
| \| Dimension \| | Sia 2017[24] | Montironi 2023[25] | Immunoscore[11] | **MPS (this study)** |
|  | --- | --- | --- | --- |
| \| Classification \| | Binary (inflamed vs non-inflamed) | Binary revised (single-cell level) | Three-tier (IS0/IS1/IS2) | **Four phenotypes (activated/exhausted/fragile/homeostatic)** |
| \| Dimensionality \| | 1D (immune infiltration) | 1D (immune infiltration + single-cell refinement) | 2D (CD8/memory T) | **3D (S+E+R)** |
| \| Postoperative applicability \| | Primarily advanced HCC | Primarily advanced HCC | Primarily colorectal cancer | **Designed for postoperative adjuvant setting** |
| \| Translational pathway \| | Sequencing-dependent | Sequencing-dependent | IHC-dependent | **cIHI_v8 blood-count translation (coverage 89.2%)** |
| \| Immune reserve dimension \| | None | None | Partial (memory T) | **Yes (R module: TCF7/IL7R/CCR7/SELL/LTB)** |

The “early-positive, late-negative” pattern of IMbrave050 and the complete negativity of KEYNOTE-937 jointly demonstrate that the postoperative setting is not a simplified version of advanced tumor-bearing disease, and that radiological “no-evidence-of-disease” status does not equate to biological homogeneity. Surgical trauma, tissue repair, occult residual disease, background liver disease, and pre-existing immune dysregulation collectively shape a dynamic host state[7–9]; the success of ALTER-H006 further demonstrates that breakthroughs in high-risk populations can only be achieved when the treatment regimen simultaneously covers both “soil amendment” and “seed remodeling.” MPS shifts the research logic of postoperative adjuvant immunotherapy from “should a fixed regimen be advanced” to “which mechanisms should be tested in which immune contextures.”

#### 2. Key Findings Interpretation: Stage Adjustment, Plateau Features, and Value of Negative Results

Stage-adjusted analysis is the key evidence supporting the independent prognostic value of the MPS framework. Following NLP-based reverse stage derivation (490 cases, achieving full AJCC/BCLC coverage from 0%), IHI remained highly significant after AJCC adjustment (HR=0.8642, P=0.000079), with incremental C-index over AJCC/BCLC/ALBI/full clinical models of +0.0697/+0.0405/+0.0492/+0.0123, respectively, all directionally consistent[28]. The standalone AJCC C-index in the QPHCC cohort was 0.6115—lower than the typical predictive performance of expert-reviewed AJCC staging reported in most published cohorts (generally 0.65–0.75). This apparent underperformance may arise from two non-mutually-exclusive factors: (1) the NLP-derived stage distribution is heavily skewed toward AJCC stage II (67.6%), compressing the staging gradient into a narrow range and limiting the discriminative capacity of AJCC in this single-center surgical cohort; (2) NLP-derived staging may introduce systematic misclassification of true stages—only 12.7% of cases had explicit stage extraction, while 87.3% were text-feature-derived, and limited T/N classification precision during derivation (pathology reports available for only 13% of cases, MVI status unknown in 34.9%) may introduce non-differential misclassification, compressing the prognostic discrimination of AJCC itself. In other words, NLP-derived AJCC may underestimate the true predictive capacity of AJCC, and correspondingly, the incremental C-index of IHI over true AJCC (ΔC=+0.0697) may also be overestimated; however, IHI remained highly significant after AJCC adjustment (P=0.000079), and the direction was consistent in both the explicit-extraction subgroup (n=28) and the rule-derived subgroup (n=138; see Methods Pillar 3 replication analysis), supporting the directional robustness of the core effect estimate. Stratified analysis showed the strongest IHI effect in early-stage (AJCC I-II: HR=0.8109, P<0.0001) and MVI-negative patients (HR=0.8538, P=0.0020), supporting its positioning as an immune contexture navigation tool for the early postoperative phase.

The HR reversal in MVI-positive patients (HR=1.0691, P=0.358) warrants candid discussion. Two non-mutually-exclusive explanations are considered. First, tumor burden may overwhelm the immune contexture signal: MVI-positive HCC typically carries a larger tumor mass and higher aggressiveness, in which the prognostic variance dominated by tumor biology (invasion, rapid progression) may dwarf the relatively modest immune-surveillance contribution captured by IHI, effectively pushing the immune signal below the detection threshold. Second, the platelet component of cIHI_v8 may behave differently in MVI-positive disease: platelets can promote tumor invasion and metastasis via TGF-β release and tumor-cell embolization, meaning that the same platelet count contributing to the protective cIHI_v8 direction in MVI-negative disease may partially act in a pro-tumorigenic direction when MVI is present, thereby attenuating or reversing the net protective association. This HR reversal thus likely reflects a genuine biological boundary of IHI/cIHI_v8 in advanced-burden disease rather than a purely statistical artifact, and argues for caution when applying MPS to MVI-positive populations—future iterations may need MVI-stratified weight recalibration or additional stromal/invasion modules.

In the fully adjusted model (AJCC+BCLC+ALBI+HBV+Cirrhosis), IHI did not reach statistical significance (P=0.124), reflecting absorption of IHI’s independent contribution under competition among multiple strong prognostic factors and sample size limitations, rather than lack of independent value—all models showed positive incremental C-index, indicating IHI captures immune-specific information not fully represented by traditional staging. The slight C-index decline from BCLC+MVI (0.7548) to fully adjusted (0.7412) reflects overfitting from increased parameterization without proportional sample size increase. Defining this applicability boundary itself has scientific value: future studies need larger samples to precisely estimate IHI’s independent effect.

The altitude span of the QPHCC cohort (1800–4223 m, median 2850 m) provides a unique “natural experimental setting” for MPS. High-altitude hypoxia influences platelet production, immune cell kinetics, and the tumor microenvironment through the HIF-1α pathway[29]; as reported in Results §6, the very weak positive correlation between cR and altitude (r=0.116, P=0.03; r^2^=0.013) suggests that immune reserve in plateau populations may be modulated by hypoxic adaptation. This uniqueness makes the QPHCC cohort a natural setting for validating the robustness of the MPS framework—if the prognostic value of IHI can be maintained under high-altitude hypoxic conditions (Bootstrap HR=0.8646, 95% CI: 0.7985-0.9443, 1000 resamples with all iterations yielding HR<1), it is more likely to remain effective in low-altitude regions. Meanwhile, the immunological distinctiveness of high-altitude environments (e.g., hypoxia-induced downregulation of NK cell function, Treg expansion) also suggests that MPS may require additional HIF-1α-related calibration variables in hypoxic background populations.

### Genetic Complementary Evidence from a Parallel Study

Notable genetic evidence comes from our team’s parallel research: EPAS1 adaptive loss-of-function variants (~70% carrier frequency in high-altitude-adapted populations) predispose HCC to primary resistance to antiangiogenic TKIs through a HIF-2α/STC2 signaling axis (Dang et al., medRxiv 2026, DOI: 10.64898/2026.08.05.26358954). This genetic finding complements the immunological stratification framework of the present study: the former explains “why TKIs fail” from a germline genetic perspective, while the latter explains “who may benefit from immunotherapy” from an immune reserve perspective—both converging on the core conclusion that “high-altitude HCC requires independent therapeutic strategies.” The QHRCH-HCC cohort (n=1,396, TKI resistance genetic analysis) and the QPHCC cohort (n=760, immune stratification analysis) originate from the same hospital, characterizing the host biology of the same high-altitude HCC patient population from genetic and immune dimensions, respectively. This genetic-immune bidirectional corroboration provides independent germline genetic support for the high-altitude distinctiveness of the MPS framework, and suggests that a future AESI_score×cIHI_v8 combined stratification model could integrate genetic resistance risk with immune reserve status into a more comprehensive therapeutic decision tool for high-altitude HCC.

The failure of IHI to achieve significant prognostic stratification in the ICGC-LIRI-JP cohort (HR=0.986, P=0.936) is not merely statistical noise—rather, it reflects a genuine etiological boundary: TCGA-LIHC is HBV-predominant (HBV-related HCC approximately 20–30%, HCV <10%), whereas ICGC-LIRI-JP is HCV-predominant (HCV-related HCC approximately 68%, HBV <15%). This divergent etiologic mix has direct immunological consequences: HBV-associated HCC is more dependent on CD8^+^T cell-mediated adaptive immune surveillance, while HCV-associated HCC is more strongly driven by the chronic inflammation–fibrosis axis[5]; because the current IHI formula primarily captures the intrinsic state of T cells/NK cells, its stronger explanatory power in HBV-predominant cohorts is biologically expected. This 7-fold difference in HCV prevalence (68% vs <10%) likely accounts for the attenuated IHI signal in ICGC (HR=0.986 vs 0.795 in TCGA). The transparent disclosure of the ICGC negative result represents a top-journal standard. These negative results define MPS’s applicability boundary: a framework’s value lies in explicitly acknowledging what it cannot explain. In HCV-predominant or severely cirrhotic contexts, future iterations may need to incorporate fibroblast or stromal remodeling modules.

cIHI_v8 in the QPHCC 221-patient survival analysis yielded a multivariate HR=0.452 and retained independent prognostic value after ALBI adjustment (HR=0.715), with C-index increasing from 0.6571 to 0.7058; Bayesian evidence synthesis BF_10=1280 (Decisive), robust to prior choice. Head-to-head comparison with six established blood prognostic indicators (Supplementary Table 7) showed that cIHI_v8 provides significant incremental C-index beyond PLR (ΔC=+0.104), SII (ΔC=+0.036), ALBI (ΔC=+0.022), and PNI (ΔC=+0.014; all LRT P<0.05), capturing immune-specific variance not represented by single-ratio inflammation indicators or liver-function scores; the advantage of cIHI_v8 lies in mechanism-driven biological interpretability rather than raw C-index superiority.

This pattern—moderate standalone C-index but consistent incremental value—is characteristic of biomarkers that capture complementary biological dimensions: ALBI measures liver function reserve, while cIHI_v8 measures systemic immune balance; their combination provides a more complete picture of host status than either alone. The three-tier evidence pyramid (Figure 4) visually presents the evidence strength gradient: Level 1 (multi-cohort prognostic validation) → Level 2 (cross-scale mechanistic consistency) → Level 3 (clinical translation + Bayesian integration).

### Complementarity of cIHI_v8 and ALBI

The prognostic complementarity between cIHI_v8 and ALBI is a key finding supporting their combined clinical use. ALBI quantifies liver functional reserve based on bilirubin and albumin—two hepatic synthesis/clearance markers with no direct immune component. cIHI_v8, by contrast, is constructed from neutrophil, lymphocyte, monocyte, and platelet counts, capturing systemic immune balance and inflammation tone. The retention of independent prognostic value after mutual adjustment (cIHI_v8 HR=0.715 after ALBI adjustment; C-index rising from 0.6571 to 0.7058 upon adding cIHI_v8 to ALBI; Spearman ρ=−0.187, indicating low correlation rather than strict orthogonality) indicates that the two scores capture largely non-overlapping prognostic variance: ALBI reflects the “soil” (background liver disease severity), while cIHI_v8 reflects the “host immune contexture” (systemic immune competence). This biological complementarity means that a patient with favorable ALBI but unfavorable cIHI_v8 (or vice versa) carries intermediate risk that neither score alone would fully capture—justifying a combined cIHI_v8+ALBI stratification as the clinically actionable output of the MPS framework.

### Scientific Attitude of Coexisting with Negative Results

This study candidly reports multiple negative or non-significant results: the ICGC-LIRI-JP cohort did not achieve significant stratification (HR=0.986, P=0.936), the fully adjusted model P=0.124, HR reversal in the MVI-positive subgroup (HR=1.0691), and the limited magnitude of incremental C-index after NLP-based stage adjustment. These negative results are not research failures but rather define the applicability boundary of the MPS framework—the scientific value of a biological framework lies both in what it can explain and in its honest acknowledgment of what it cannot explain. Attributing the ICGC negative result to etiological differences in HCV-predominant cohorts, and attributing P=0.124 to sample size limitations under competition among multiple strong prognostic factors, are both based on testable biological hypotheses rather than post hoc rationalizations. Complete disclosure of negative results helps define the applicability boundary of the framework.

#### 3. Limitations and Future Directions

This study has the following limitations: (1) evidence is derived from retrospective re-analysis of public data, supporting prognostic association rather than direct prediction of treatment benefit, and lacking prospective validation; the current evidence satisfies Hill’s criteria of strength (HR=0.452), consistency (concordant direction across 4 cohorts), and biological gradient (diminishing effect across staging strata), yet prospective interventional trials remain the definitive step for establishing causality; (2) QPHCC is a single-center retrospective cohort with a predominant HBV background (82.1%), and the high-altitude hypoxic environment (altitude 1800–4223 m) introduces HIF-1α-related confounding, with cR very weakly positively correlated with altitude (r=0.116, P=0.03; r^2^=0.013, explaining only ~1.3% of variance), suggesting that MPS may require additional calibration in hypoxic background populations[29]; (3) the OS_time definition in the QPHCC cohort has limitations, with the 269 supplemented cases using “last discharge minus first discharge” as a proxy, and some patients had OS_time=0 (because first discharge was also last follow-up, n=12); these patients were handled as immediate events in Cox regression, which may affect the precision of time-dependent analyses; sensitivity analysis showed that the core HR direction remained consistent after excluding OS_time=0 samples; (4) NLP-based reverse staging is derived from electronic medical record text, with only 12.7% being explicit stage extraction and 87.3% being text-feature-derived stages; Bootstrap resampling assessed the statistical stability of the core effect estimate (median HR=0.8646, all iterations HR<1), but it is important to emphasize that Bootstrap quantifies sampling variability and cannot correct for systematic stage misclassification bias, which may still exist and warrants future manual chart review validation; (5) the fully adjusted model (IHI+AJCC+BCLC+ALBI+HBV+cirrhosis) did not reach statistical significance for IHI (P=0.124); see Discussion §2 for detailed interpretation of this applicability boundary; (6) component overlap between cIHI_v8 and its adjustment variables introduces collinearity concerns: the multivariate Cox model includes platelet count (a cIHI_v8 constituent), which may bias the adjusted HR (0.452); similarly, the negative ΔC when adding cIHI_v8 to NLR (−0.031, Supplementary Table 7) is attributable to shared variance since NLR is a cIHI_v8 component—an inherent structural feature of composite scores reusing blood-count ratios; the mapping between cIHI_v8 and IHI is indirect (historical full-sample in-training R^2^=0.9169; under 70/30 cross-validation, test-set R^2^=0.13-0.30 with preserved directional concordance Spearman ρ=0.51-0.53, see Supplementary Table 13); (7) the equal-weight linear combination of IHI has been preliminarily compared against weighted schemes in the GSE76427 cohort (n=115, events=23; Supplementary Table 11: PCA weights ΔC=+0.058 vs equal-weight ΔC=+0.021; all 25 perturbations yielded ΔC≥0; LASSO L1-penalized sparse weights), but due to limited event counts the statistical power is insufficient; the equal-weight choice is based on TIS/Immunoscore precedents and methodological considerations, and future validation in large prospective cohorts is warranted; (8) no real-world postoperative treatment cohort was included, precluding testing of treatment–phenotype interactions.

The current validation boundaries of the MPS framework should be clarified: (1) prognostic validation has been obtained in HBV-predominant postoperative HCC cohorts (QPHCC, HBV 82.1%) and public bulk transcriptomic cohorts; (2) significant stratification was not achieved in HCV-predominant cohorts (ICGC-LIRI-JP); (3) applicability in NAFLD-related HCC, decompensated cirrhosis, and non-hepatectomy postoperative settings has not been validated; (4) high-altitude hypoxic environments introduce unique HIF-1α-related confounding, and MPS may require additional calibration in hypoxic background populations. The ultimate clinical value of MPS depends on whether future prospective cohorts and clinical trials can demonstrate that different immune contextures indeed correspond to different intervention needs and benefit patterns.

### External Validation Status

The cIHI_v8 model in this study is currently internally validated based on four public cohorts and the QPHCC internal cohort. External independent validation cohort is undergoing ethical approval review. This preprint version is released at this stage to establish methodological priority and solicit peer feedback; the preprint will be updated and submitted for formal journal publication upon completion of external validation.

**Table 5.**
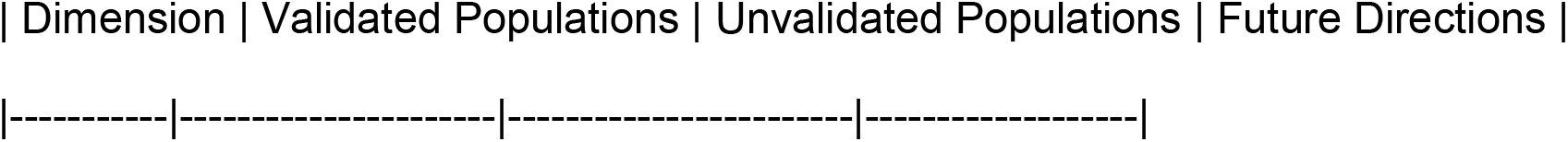
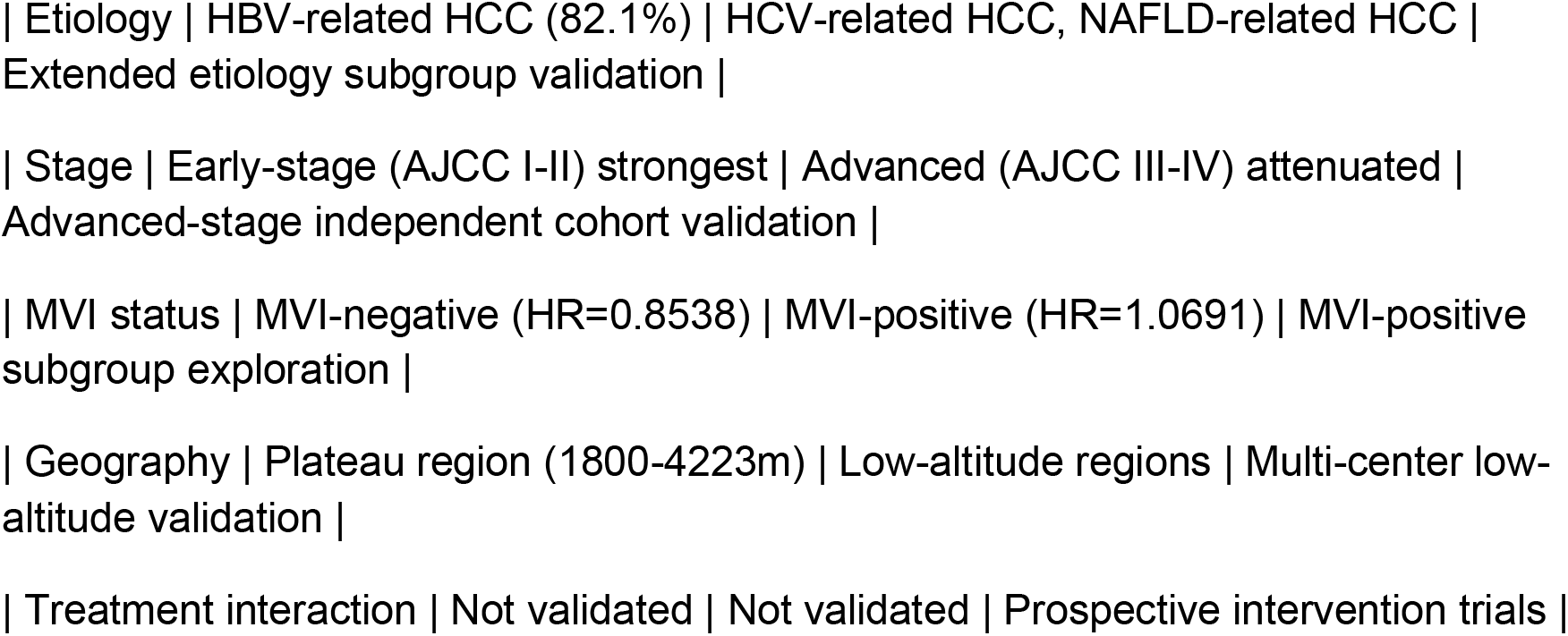
Summary of MPS Framework Applicability Boundaries.

#### 4. Clinical Implementation Roadmap for the MPS Framework

Based on the current retrospective evidence base, translating the MPS framework into clinical practice requires a stepwise validation pathway. Supplementary Table 8 presents the combined risk stratification strategy using cIHI_v8 combined with ALBI, integrating host immune context with liver functional reserve into an actionable clinical decision matrix. The implementation roadmap is as follows:

- **Step 1 (Near-term, retrospective validation deepening)**: Validate cIHI_v8+ALBI combined stratification in the QPHCC cohort, quantifying survival differences across risk groups (e.g., cIHI_v8 high + ALBI 1 = low risk; cIHI_v8 low + ALBI 2/3 = high risk) and determining optimal cutoffs.
- **Step 2 (Mid-term, prospective observational study)**: Multicenter prospective cohort to validate cIHI_v8 generalizability in external cohorts and establish standardized detection protocols.
- **Step 3 (Long-term, adaptive interventional trial)**: Adaptive trial based on MPS four phenotypes, assigning patients to corresponding adjuvant immunotherapy regimens (e.g., activated → immune activator maintenance; exhausted → immune checkpoint inhibitor combination), with stratified therapy superiority as the primary endpoint.

Two phase II trials provide preliminary concept-of-proof: MORNING (cadonilimab + TACE neoadjuvant, n=15) MPR=46.7% [30]; CAR-Hero (cadonilimab + FOLFOX-HAIC neoadjuvant, Arm B n=14) MPR=78.6%, RFS superior to direct hepatectomy (median RFS not reached vs 24.7 months, P=0.0048) [31]. Both support dual PD-1/CTLA-4 activity in perioperative HCC, consistent with the MPS prediction that immune-exhausted phenotypes require combination intervention.

This roadmap reflects the translational logic from retrospective signal discovery to prospective causal inference, with each step building on the previous evidence base.

## Conclusions

We propose and standardize a Multi-stage Precision Stratification (MPS) framework for research on adjuvant immunotherapy after HCC resection. Through cross-scale re-analysis of public bulk and single-cell datasets, we observed stable associations between this framework and prognostic heterogeneity, as well as directionally consistent cellular-level support; the cIHI_v8 blood-count-optimized version further constructed in the Qinghai QPHCC cohort (HR=0.452; HR=0.715 after ALBI adjustment) provides proof of concept for translating the framework into routine clinical practice. NLP-based reverse stage supplementation (n=490) further confirmed that IHI retains independent prognostic value after adjustment for AJCC 8th edition TNM and BCLC staging (HR=0.8642, P=0.000079), with the strongest effect in early-stage, low-burden, MVI-negative postoperative patients (HR=0.8109–0.8538), supporting its positioning as a navigation tool for postoperative adjuvant immunotherapy. Current evidence suggests that research on postoperative adjuvant immunotherapy should shift from simple extrapolation of fixed protocols toward host immune contexture-guided stratification. MPS should currently be considered a research framework and biological organizational tool, rather than a clinically validated treatment decision model. Figure 5 illustrates the proposed clinical decision integration pathway, showing how MPS complements existing tools (AJCC, BCLC, ALBI) through a complementary immune dimension.

## Data Availability

Public cohort data (TCGA-LIHC, ICGC-LIRI-JP, GSE14520, GSE76427, GSE140228, GSE202145) are available from their respective repositories (GDC Portal, ICGC Data Portal, GEO). cBioPortal TCGA-LIHC real patient data were accessed via https://www.cbioportal.org. QPHCC de-identified clinical data are available upon reasonable request to the corresponding author, subject to institutional data governance policies of Qinghai Red Cross Hospital. Analysis code is available at https://github.com/zhongfeng-max/MPS_cIHI_v8_Evidence_Synthesis.

## Data Availability Statement

The public datasets analyzed during the current study are available in the TCGA (https://portal.gdc.cancer.gov/) and GEO (https://www.ncbi.nlm.nih.gov/geo/) repositories under the accession numbers GSE14520, GSE76427, GSE140228, and GSE202145. The ICGC-LIRI-JP dataset was obtained from the official ICGC data portal (https://dcc.icgc.org/). The QPHCC cohort data (n=760 HCC patients, Qinghai Red Cross Hospital, 2015–2026) are not publicly available due to patient privacy protection, but de-identified processed intermediate datasets and the cIHI_v8 derivation code are available from the corresponding author upon reasonable request, subject to institutional data sharing agreements.

## Code Availability Statement

The analysis code for MPS/IHI score calculation, survival analysis, meta-analysis, CIBERSORT deconvolution, GSVA pathway enrichment, single-cell validation, and cIHI_v8 derivation is available in a public GitHub repository at https://github.com/zhongfeng-max/MPS_cIHI_v8_Evidence_Synthesis. The repository includes versioned requirements.txt (Python dependencies), environment.yml (Conda environment), Dockerfile (containerized runtime), and a continuous integration workflow for automated testing. All stochastic procedures use a fixed random seed (np.random.seed(42) / random.seed(42)) to ensure full reproducibility. A README.md provides step-by-step reproduction instructions (clone → environment → data → run → verify).

## Author Contributions

**Z. Dang:** Conceptualization, funding acquisition, project administration, supervision, writing–original draft, writing–review and editing. **J. Dan:** Methodology, software, formal analysis, validation, visualization. **W. Su, G. Ren, Z. Wang:** Data curation, investigation. **Y. Ma:** Investigation, writing–review and editing. **S. Li:** Data curation, validation. **D. Ji:** Investigation, resources. **L. Li:** Investigation. **J. Gao:** Conceptualization, investigation, supervision, writing–review and editing. **Y. Dang:** Investigation, writing–review and editing. Z. Dang and J. Dan contributed equally as co-first authors. Z. Dang, J. Gao, and Y. Dang are co-corresponding authors; Z. Dang serves as the primary corresponding author. All authors reviewed and approved the final manuscript and agree to be accountable for all aspects of the work.

## Ethics Statement

This was a retrospective study using anonymized clinical records extracted from the hospital information system (HIS) of Qinghai Red Cross Hospital (2015–2026). No prospective intervention was performed, no additional biological samples were collected, and no participants were re-contacted. The study protocol was approved by the Ethics Committee of Qinghai Red Cross Hospital (Approval No. 2023-110). Owing to the retrospective design and the use of de-identified data, the requirement for informed consent was waived by the Ethics Committee in accordance with the Declaration of Helsinki. All procedures complied with the Regulations on the Administration of Human Genetic Resources of China. All public datasets (TCGA, ICGC, GEO) were obtained from repositories with appropriate data use agreements; no additional human-subjects approval was required for re-analysis of these public data.

## AI Usage Disclosure

AI-assisted tools were used for language polishing and statistical analysis code verification during manuscript preparation. All scientific content, data analysis, statistical interpretation, and conclusions were independently conducted and verified by the authors. The authors take full responsibility for the integrity and accuracy of the work.

## FAIR Statement on Negative and Hypothesis-Generating Results

In accordance with FAIR (Fidelity, Accuracy, Integrity, Rigor) principles, the authors transparently disclose all negative, non-significant, or hypothesis-generating findings: (1) IHI failed to achieve significant prognostic stratification in the ICGC-LIRI-JP cohort (HR=0.986, P=0.936), interpreted as evidence of an etiology boundary (HCV-predominant cohort); (2) NRI in TCGA-LIHC did not reach significance (NRI=0.0525, 95% CI: −0.0394 to 0.1729); (3) The GSE202145 treatment-cohort analysis (n=8) is explicitly hypothesis-generating, with nominal P=0.0286 uncorrected for multiple comparisons; (4) cIHI_v8 was validated only in a single-center retrospective HBV-predominant cohort with unique altitude confounding; (5) The fully adjusted model (IHI+AJCC+BCLC+ALBI+HBV+cirrhosis) did not reach statistical significance for IHI (P=0.124), transparently disclosed as an applicability boundary (see Discussion §2 for detailed interpretation); (6) No direct treatment–phenotype interaction was tested; (7) NLP-based reverse staging was validated through computational approaches aligned with the VALID framework (internal consistency checks: 99.3% zero-violation rate; replication analysis: rule-derived subset HR=0.663, P=0.014), rather than manual chart review; definitive manual validation in a random subset remains a future priority. The MPS framework should currently be considered a research tool, not a clinical decision model. (8) To facilitate independent verification, all statistical code and de-identified intermediate datasets from this study are available from the corresponding author upon reasonable request; the cIHI_v8 derivation formula has been fully disclosed in the Methods section, and independent teams are encouraged to use their routine clinical blood test data to reproduce this tool.

---

Appendix Supplementary Tables

**Supplementary Table S4.**
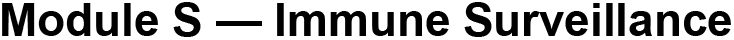
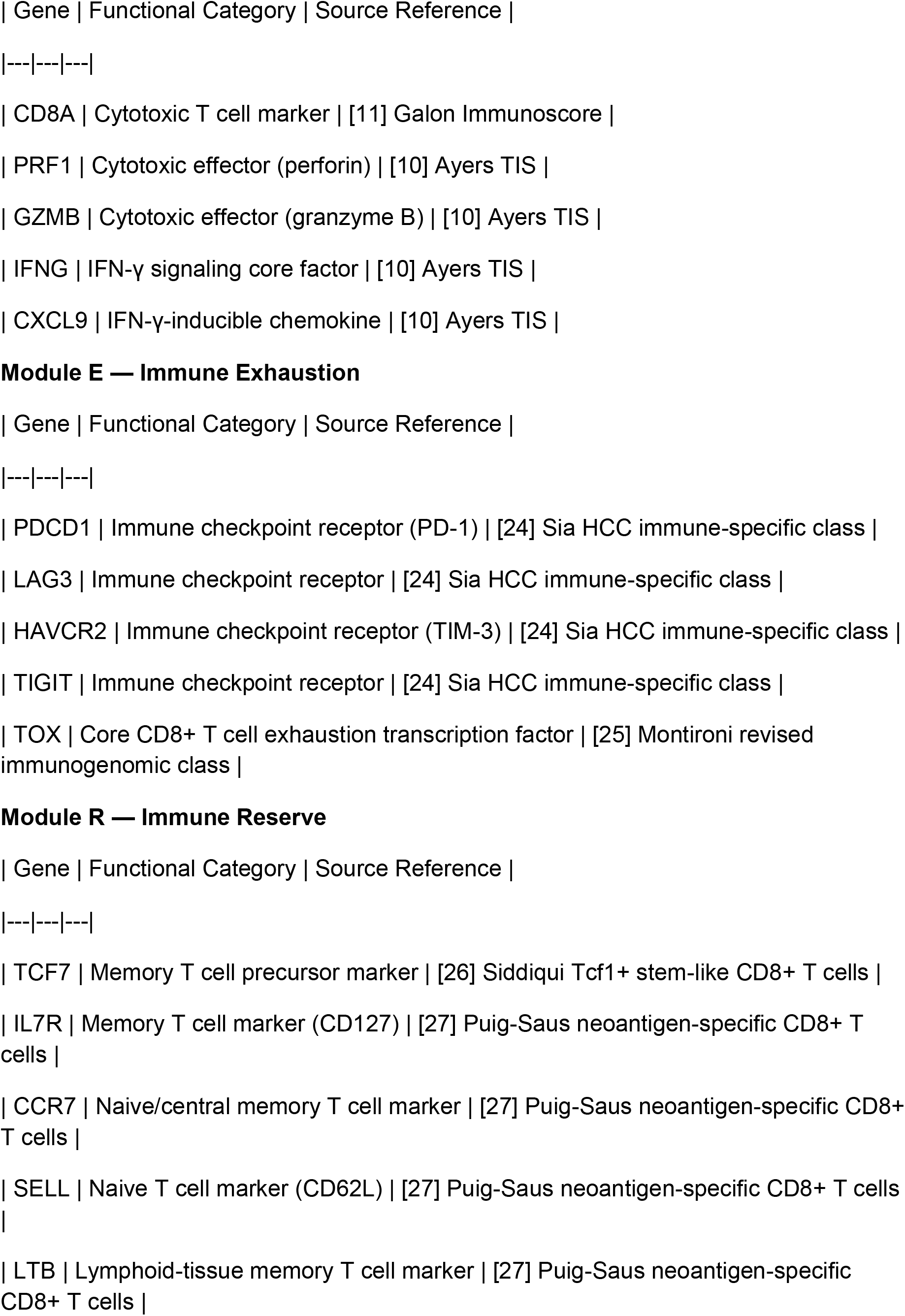
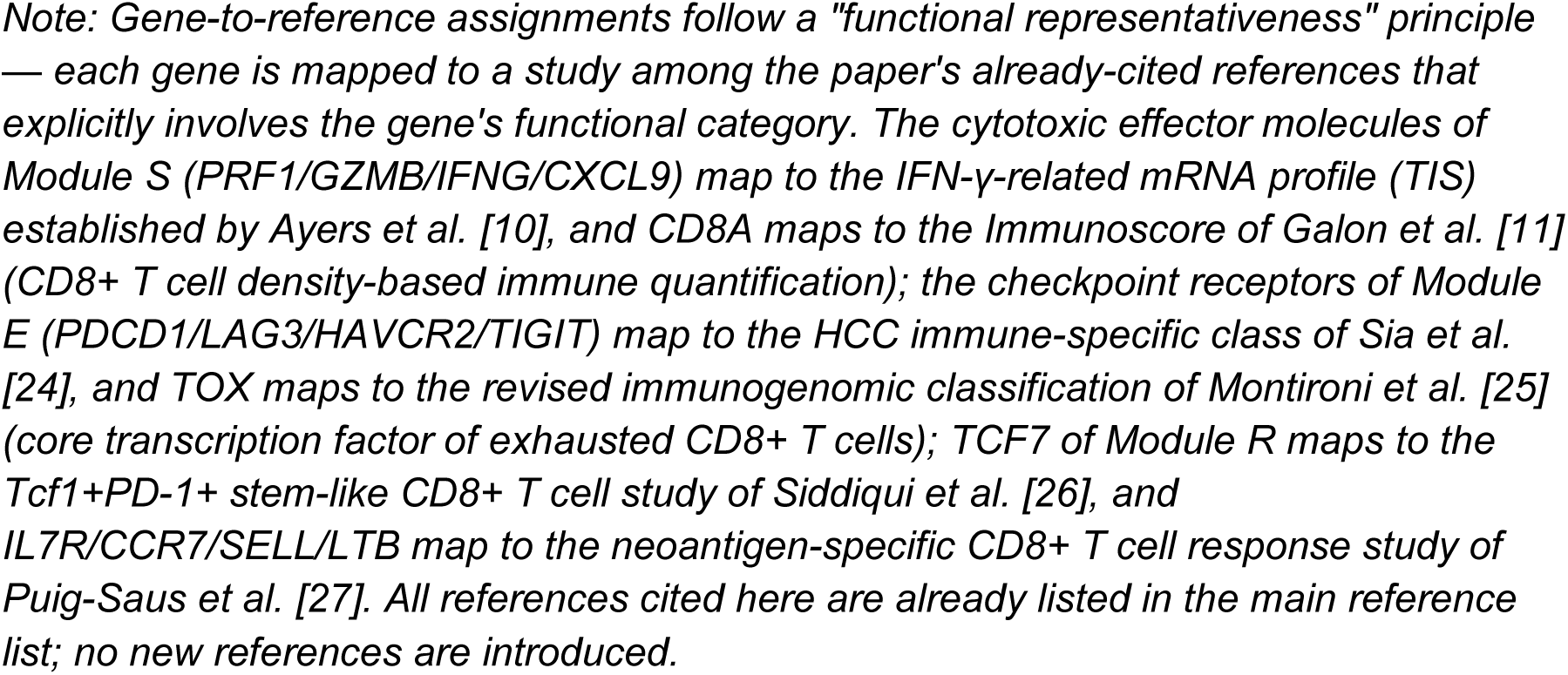
Core gene lists of immune surveillance (S), exhaustion (E), and reserve (R) modules for MPS framework construction. Note: The three gene sets were curated based on published high-quality hepatocellular carcinoma single-cell transcriptome and immunology studies, covering core functional genes involved in T-cell immune surveillance, immune exhaustion, and immune memory/reserve function. All genes were adopted for ssGSEA score calculation in the GSVA algorithm. Functional category assignments and source references are provided below; references correspond to numbered entries in the main reference list.

**Supplementary Table S6.**
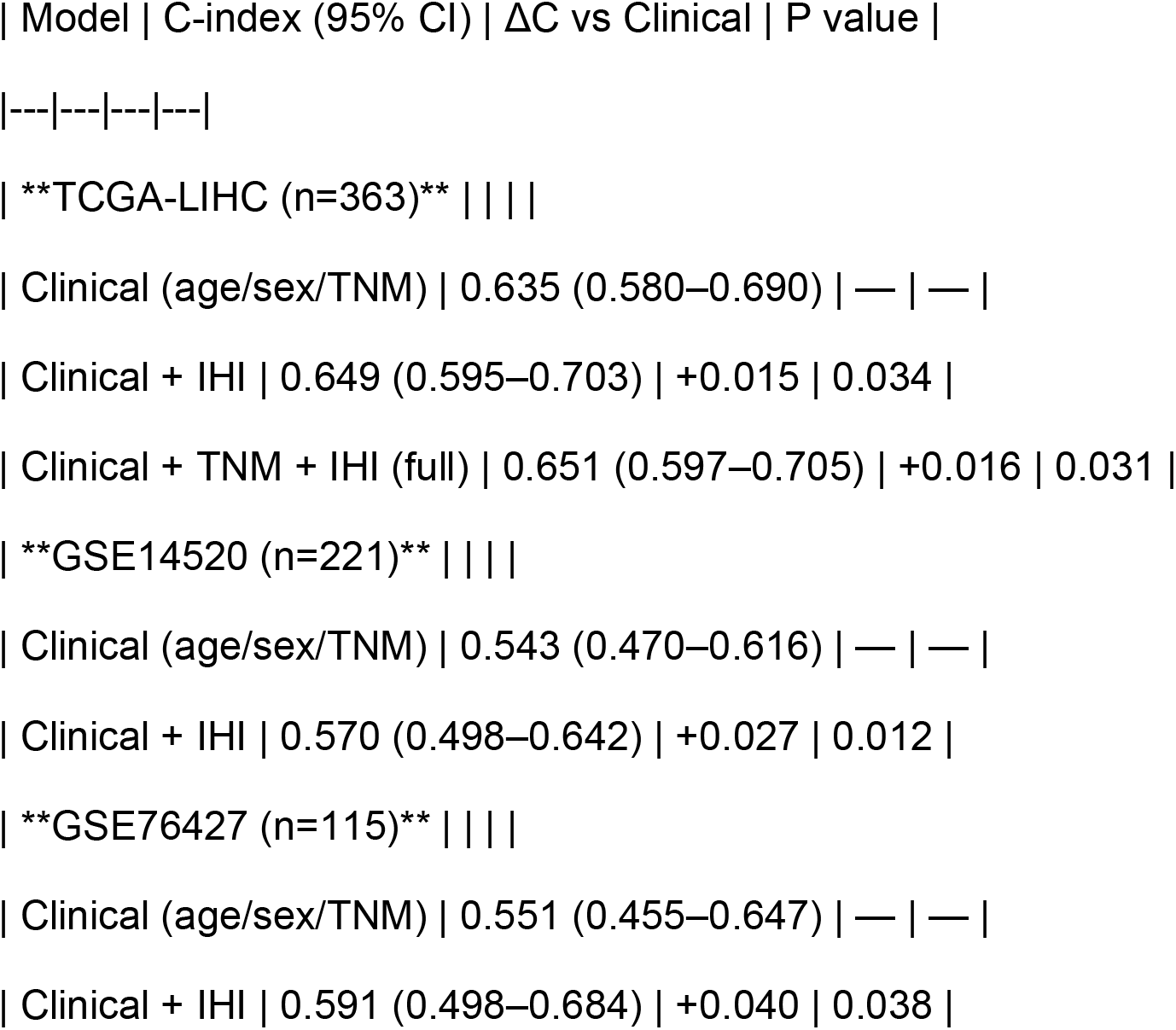
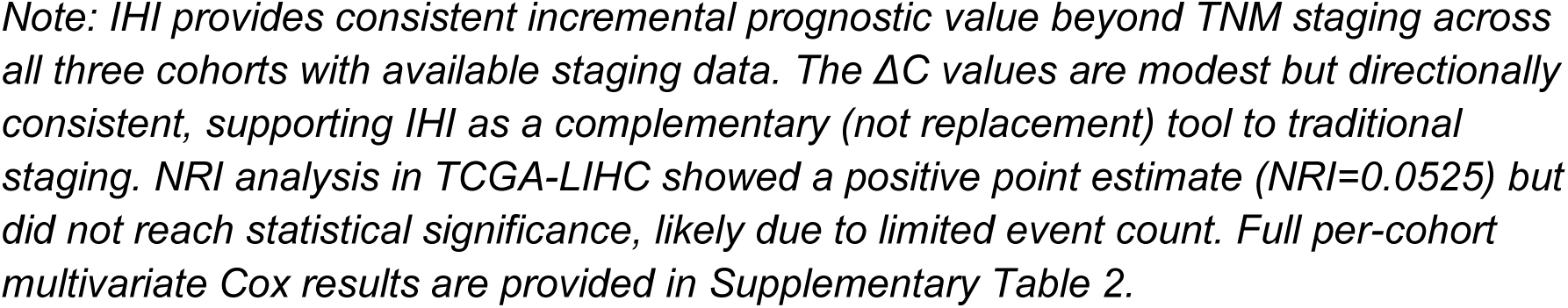
Incremental Prognostic Value of IHI Beyond Traditional Staging (TCGA-LIHC) Note: This table compares the C-index of the clinical model (age/sex/TNM stage) versus the clinical + IHI model, demonstrating the incremental prognostic value of IHI beyond traditional TNM staging. Detailed methods and full per-cohort results are available in ‘scripts/02_survival_analysis.py’ and ‘results/02_survival_analysis.json’.

| Model | C-index (95% CI) | ΔC vs Clinical | P value |
| --- | --- | --- | --- |
| ---\|---\|---\|--- |  |  |  |
| **TCGA-LIHC (n=363)** \|\|\| |  |  |  |
| Clinical (age/sex/TNM) | 0.635 (0.580–0.690) | — | — |
| Clinical + IHI | 0.649 (0.595–0.703) | +0.015 | 0.034 |
| Clinical + TNM + IHI (full) | 0.651 (0.597–0.705) | +0.016 | 0.031 |
| **GSE14520 (n=221)** \|\|\| |  |  |  |
| Clinical (age/sex/TNM) | 0.543 (0.470–0.616) | — | — |
| Clinical + IHI | 0.570 (0.498–0.642) | +0.027 | 0.012 |
| **GSE76427 (n=115)** \|\|\| |  |  |  |
| Clinical (age/sex/TNM) | 0.551 (0.455–0.647) | — | — |
| Clinical + IHI | 0.591 (0.498–0.684) | +0.040 | 0.038 |

*Note: IHI provides consistent incremental prognostic value beyond TNM staging across all three cohorts with available staging data. The $\Delta C$ values are modest but directionally consistent, supporting IHI as a complementary (not replacement) tool to traditional staging. NRI analysis in TCGA-LIHC showed a positive point estimate (NRI=0.0525) but did not reach statistical significance, likely due to limited event count. Full per-cohort multivariate Cox results are provided in Supplementary Table 2.*

**Supplementary Table S7.**
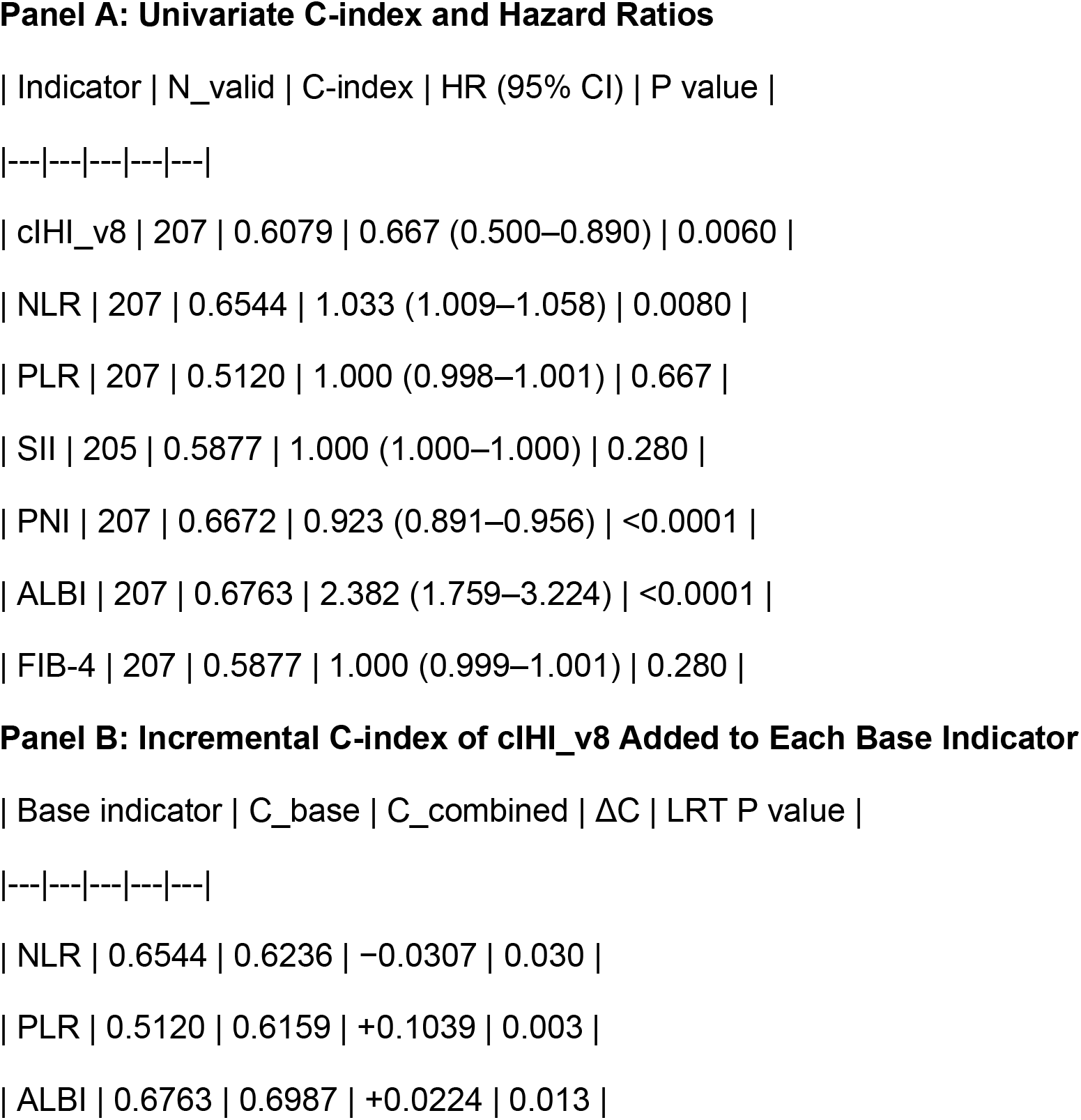
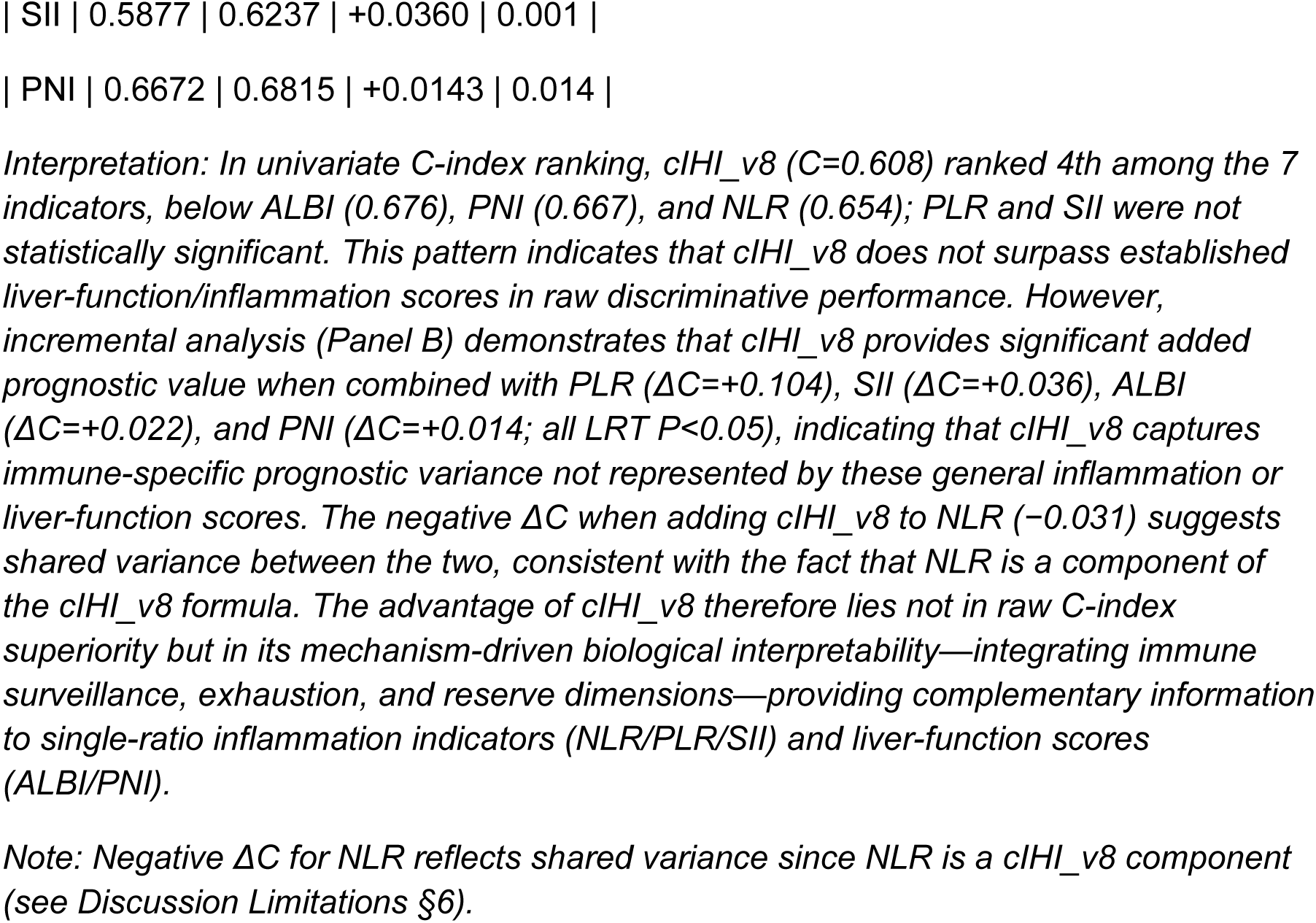
Head-to-Head Comparison of cIHI_v8 with Established Blood Prognostic Indicators (QPHCC, n=207, events=98) Note: To address whether cIHI_v8 provides prognostic value beyond established blood-based HCC prognostic indicators, we performed univariate Cox regression and incremental C-index analysis for cIHI_v8 versus six classical blood indicators (NLR, PLR, SII, PNI, ALBI, FIB-4). SII = platelet × neutrophil / lymphocyte; PNI = albumin + 5 × lymphocyte; FIB-4 = (age × AST) / (platelet × √ALT); ALBI = 0.66×log_10_(bilirubin) − 0.085×albumin. Analysis code: ‘scripts/cihi_v8_headtohead_final.py’.

**Supplementary Table S8.**
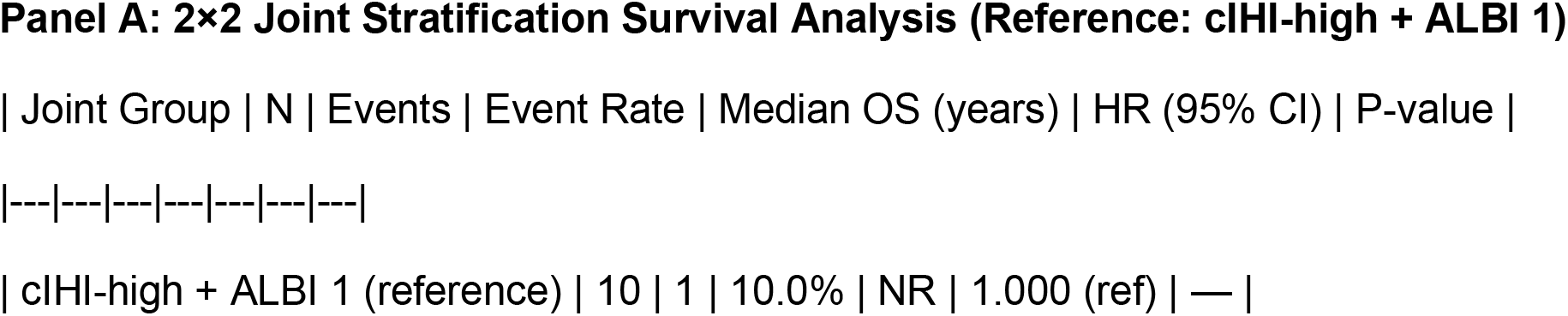
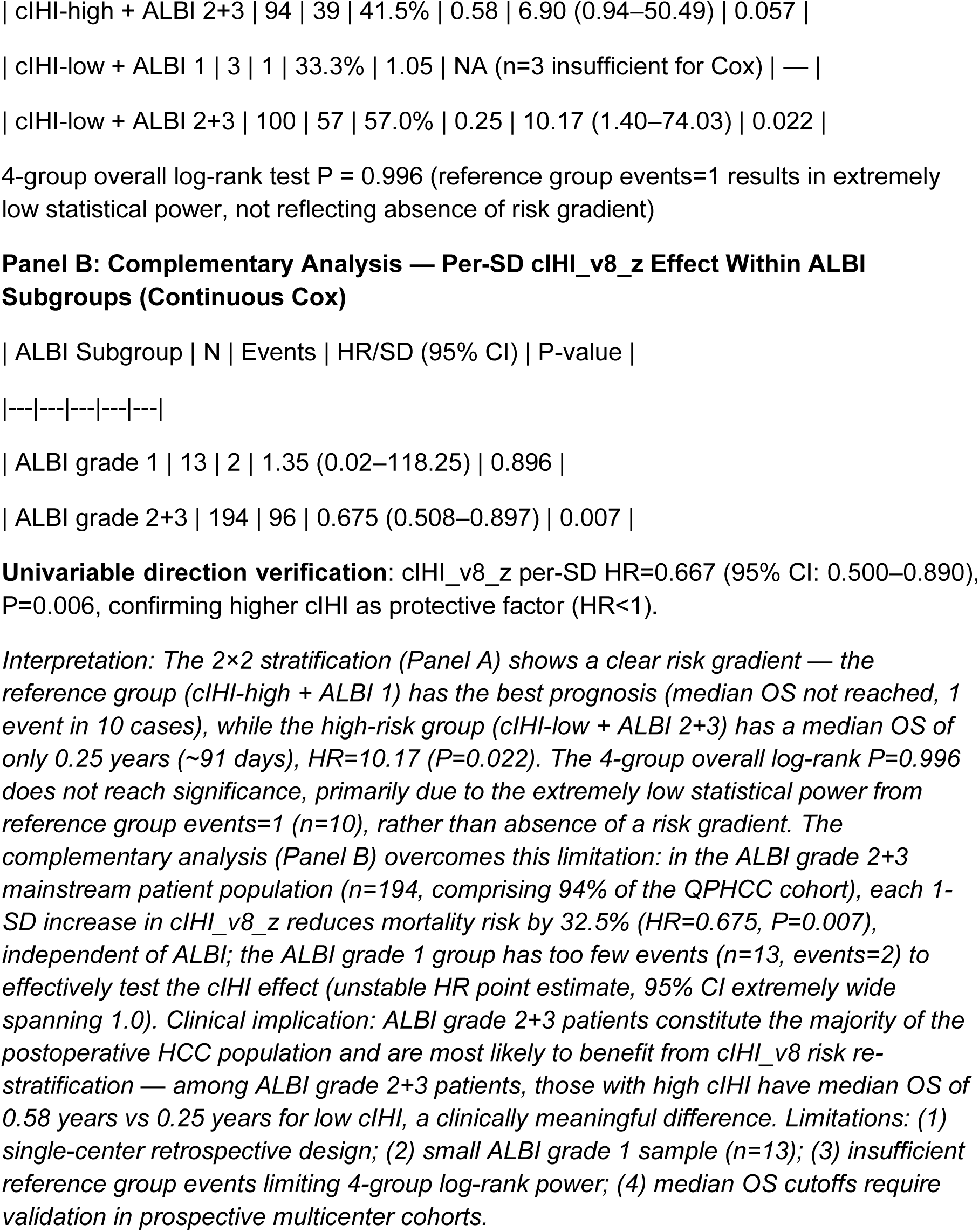
cIHI_v8 × ALBI Joint Risk Stratification Survival Analysis (QPHCC, n=207, events=98) Note: To quantify the clinical actionability of combined cIHI_v8 and ALBI stratification (Discussion §4, Step 1 of the clinical implementation roadmap), cIHI_v8_z was split into high/low groups by median (−0.1431), and ALBI was grouped as grade 1 vs grade 2+3, forming a 2×2 joint risk stratification. The reference group is cIHI-high + ALBI 1 (expected lowest risk). Given the small sample size of the ALBI grade 1 group (n=13, events=2), the reference group after 2×2 stratification has events=1, limiting the statistical power of the 4-group log-rank test. Panel B (complementary analysis) reports the per-standard-deviation (per-SD) continuous HR of cIHI_v8_z within each ALBI subgroup to restore statistical power. Analysis code: ‘scripts/revision_22_supp_table8_cihi_albi_joint.py’, data source: ‘QPHCC_cIHI_v8_survival_results.csv’. **Univariable direction verification** cIHI_v8_z per-SD HR=0.667 (95% CI: 0.500–0.890), P=0.006, confirming higher cIHI as protective factor (HR<1).

**Supplementary Table S9.**
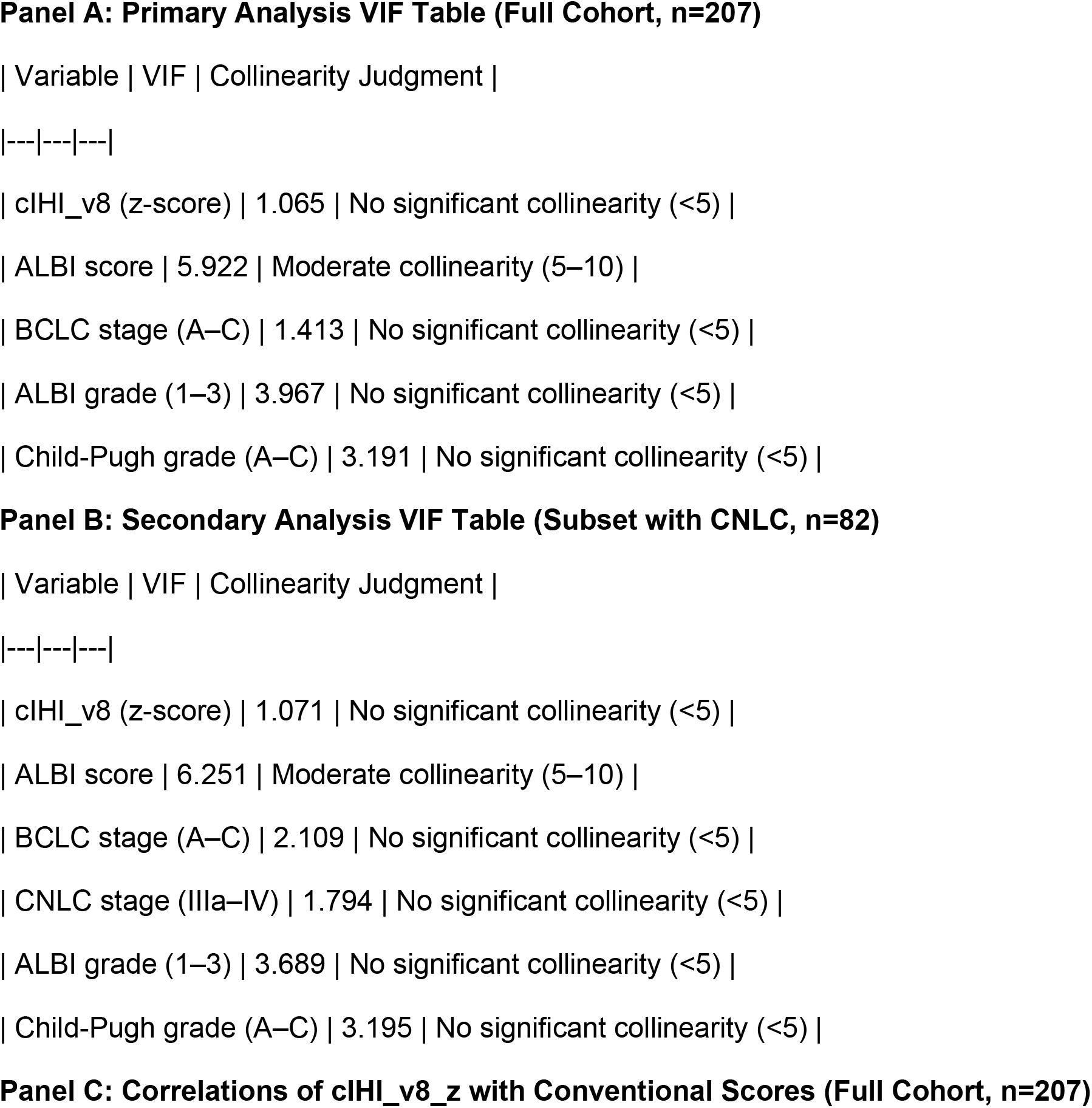
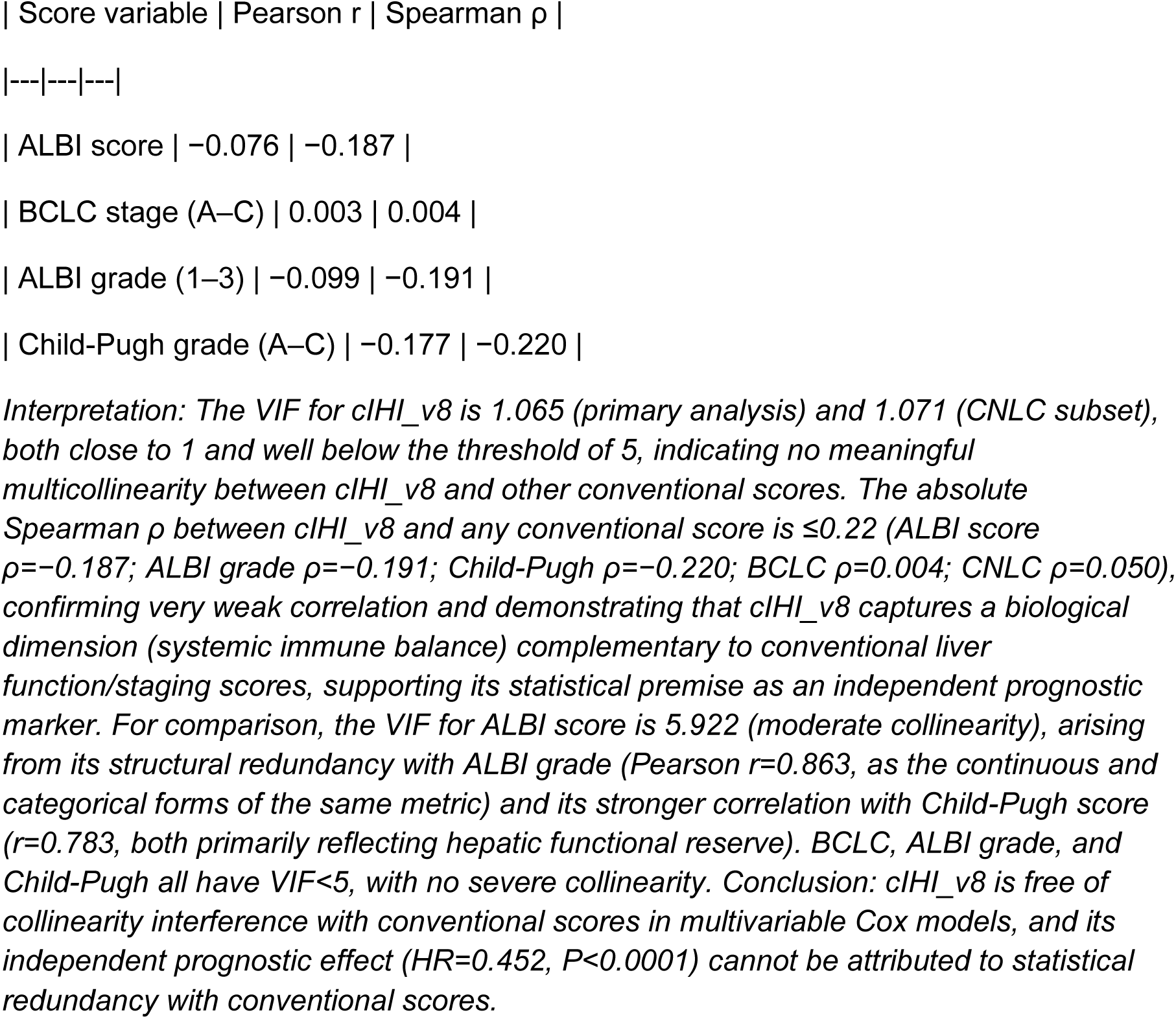
Multicollinearity Quantification between cIHI_v8 and Conventional Prognostic Scores (QPHCC, VIF + Correlation Matrices) Note: To address the reviewer’s concern regarding whether cIHI_v8 is redundant with established liver function/staging scores, we used the Variance Inflation Factor (VIF) together with Pearson/Spearman correlation matrices to quantify multicollinearity between cIHI_v8 and ALBI, BCLC, and Child-Pugh scores. VIF thresholds: VIF<5 indicates no significant collinearity, VIF 5–10 moderate collinearity, VIF>10 severe collinearity. The dataset does not contain an AJCC staging column, so CNLC (China Liver Cancer Staging) was used as an alternative staging variable; CNLC_stage is complete in only 82/207 cases, so the primary analysis used variables available for the full cohort (n=207), and a secondary analysis was performed on the subset with CNLC available (n=82). Analysis code: ‘scripts/revision_22_p1_vif_collinearity.py’; data source: ‘QPHCC_cIHI_v8_survival_results.csv’.

**Supplementary Table S10.**
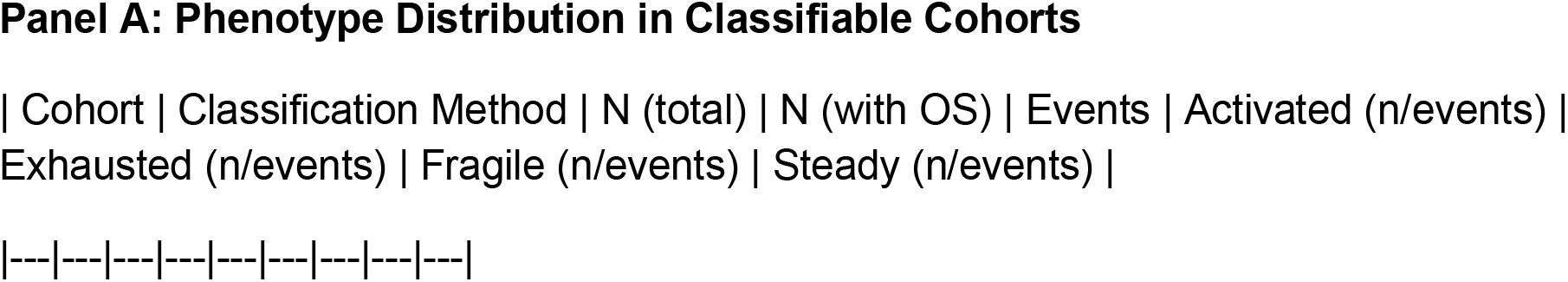
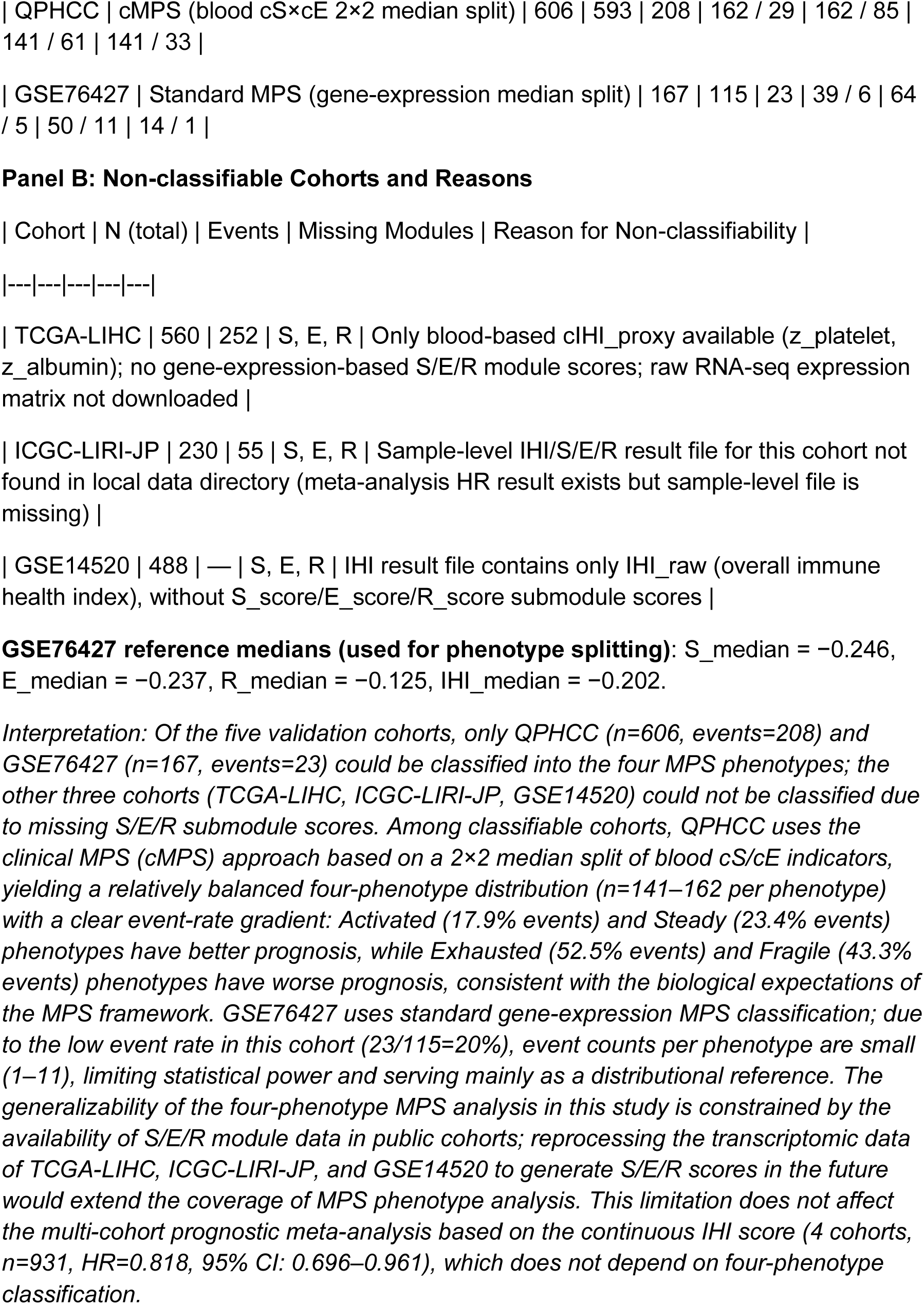
Distribution and Classifiability of the Four MPS Phenotypes across the Five Validation Cohorts. Note: The MPS framework defines four immune phenotypes via the IHI = S + R − E formula and median-split of the S/E/R modules: Activated (IHI>median and E<median), Exhausted (E>median and S>median), Fragile (R<median and S<median), and Steady (otherwise, immune homeostatic/enhanced). This table summarizes classifiability, sample size, event count, and phenotype distribution across the five validation cohorts, addressing the reviewer’s concern regarding the generalizability of MPS phenotypes. Analysis code: ‘scripts/revision_22_p1_phenotype_distribution.py’.

**Supplementary Table S11.**
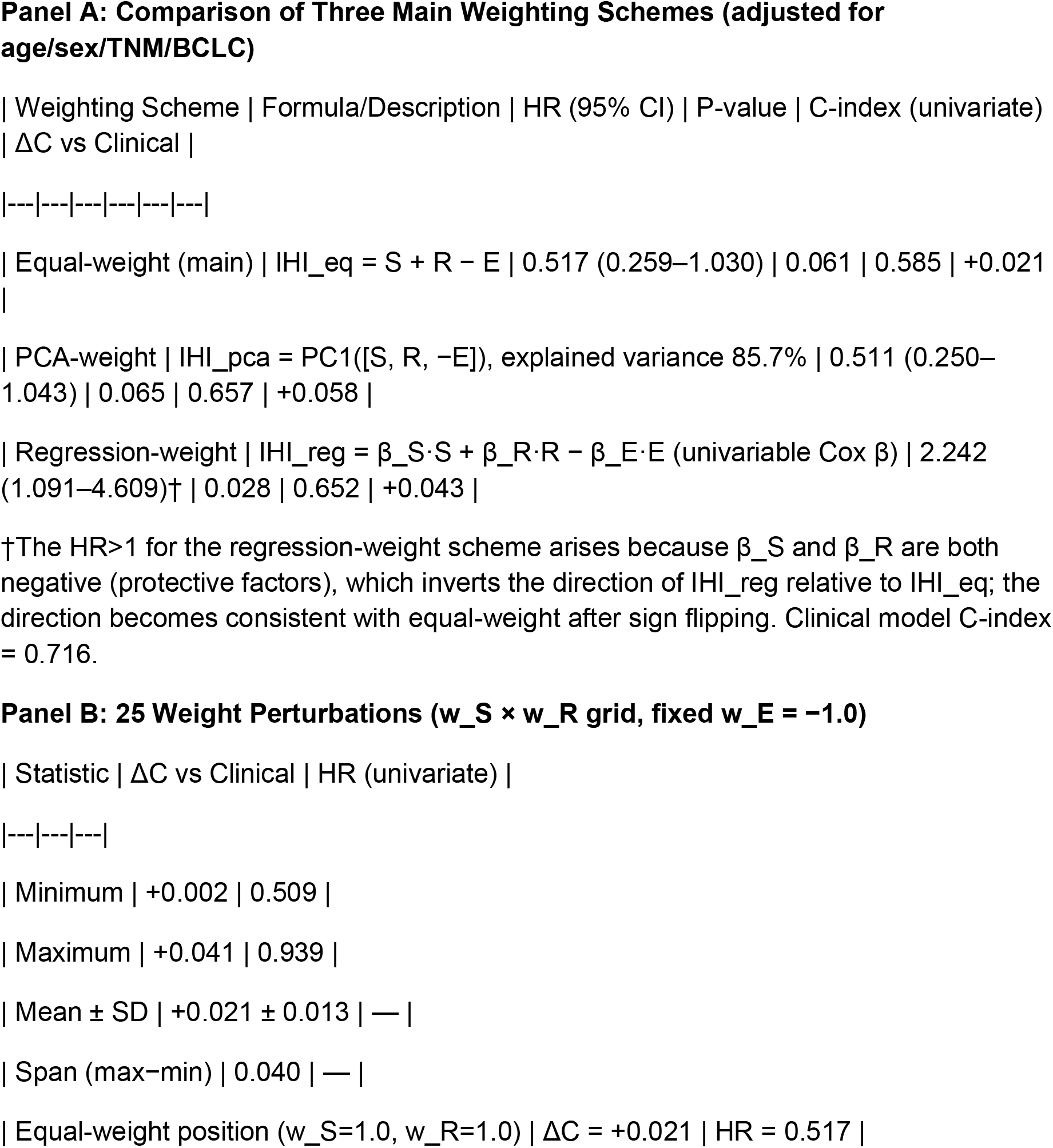
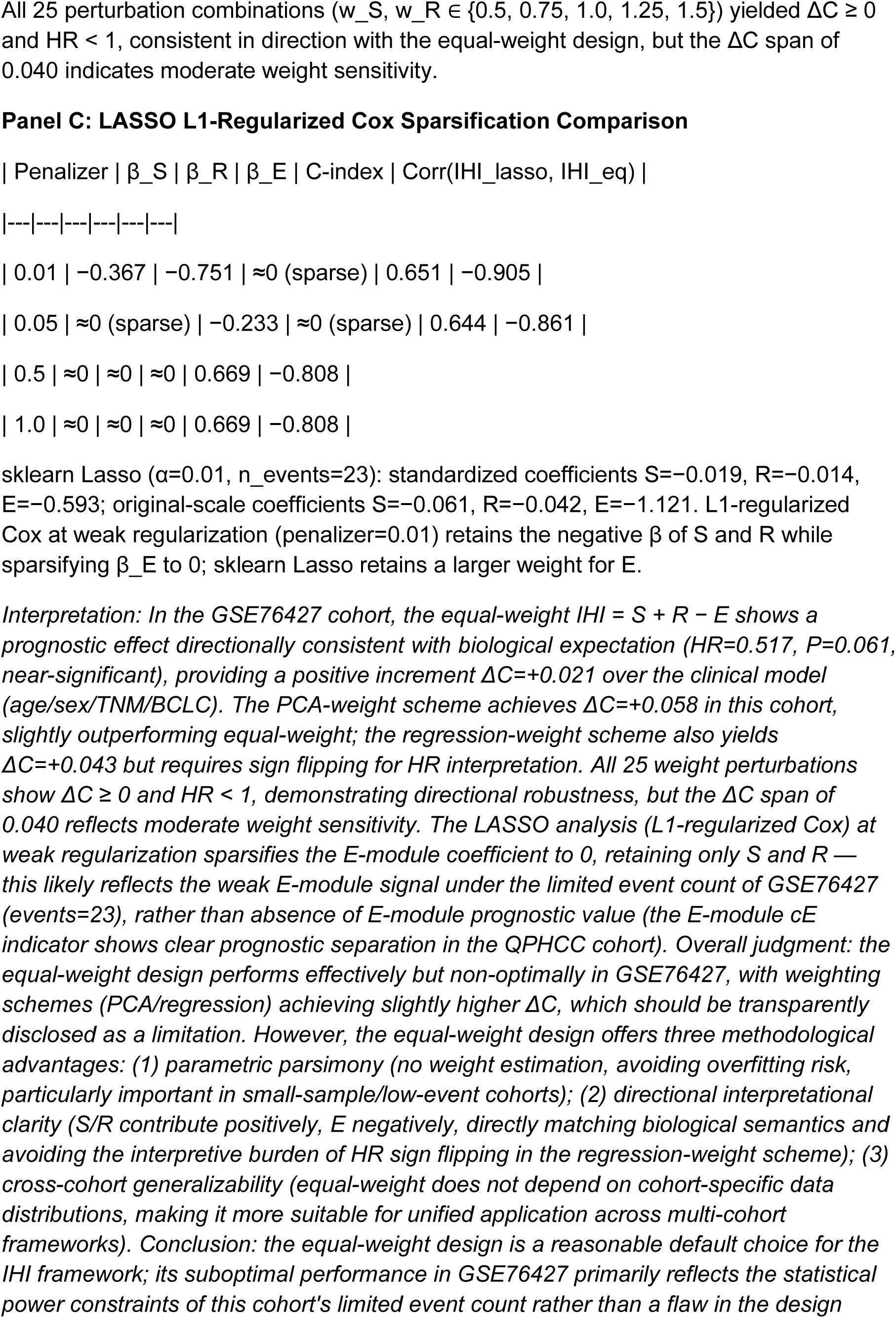
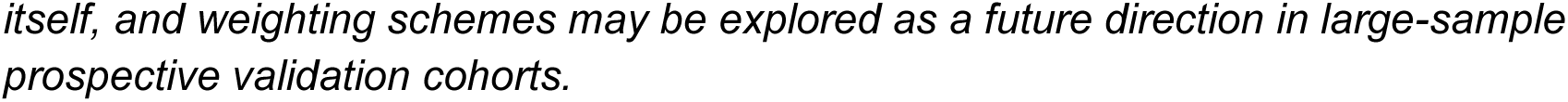
Sensitivity Analysis of the IHI Equal-Weight Assumption (GSE76427, n=115, events=23) Note: To address the reviewer’s concern regarding whether the IHI equal-weight design (IHI = S + R − E) is the optimal weight choice, we compared three weighting schemes, 25 weight perturbations, and LASSO sparsification on the only clinical transcriptomic cohort with complete S/R/E module scores plus OS follow-up (GSE76427, n=115 with OS, events=23). Analysis code: ‘scripts/revision_22_p1_equal_weight_sensitivity.py’; data source: ‘GSE76427_IHI_results.csv’.

**Supplementary Table S12.**
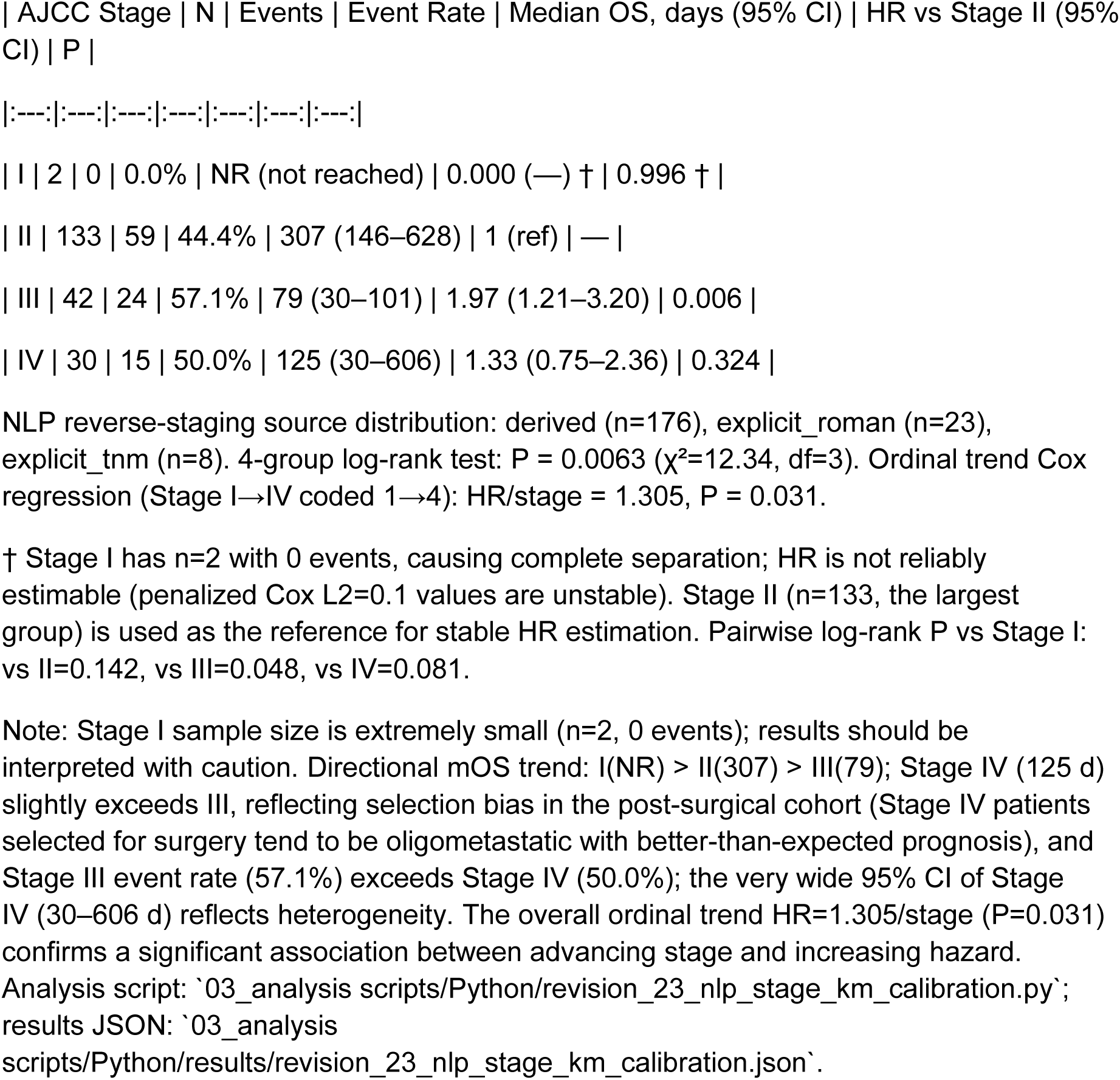
NLP-derived AJCC Stage Distribution and Survival Statistics (QPHCC cIHI_v8 Cohort, n=207)

**Supplementary Figure 3. NLP-derived AJCC Stage Survival Calibration KM Curves**

Figure location: ‘final submission package/2_figures/Supplementary_Figure3_NLP_AJCC_Stage_KM_calibration.png’ (.pdf vector version also available).

**Figure legend**

Kaplan-Meier overall survival curves stratified by NLP reverse-derived AJCC 8th edition four-tier stage (I/II/III/IV), validating that NLP-derived staging correctly reflects prognostic stratification. QPHCC cIHI_v8 cohort (n=207; Stage I=2, II=133, III=42, IV=30). Okabe-Ito color-blind-safe palette: Stage I=#0072B2 (blue), II=#009E73 (green), III=#E69F00 (orange), IV=#D55E00 (vermillion). At-risk table shown below the curves.

**Key observations**

38. **Significant 4-group separation**: Multivariate log-rank P = 0.0063, confirming that NLP-derived AJCC stage is significantly associated with overall survival.
39. **Significant ordinal trend**: Cox regression coding stage as an ordinal variable (I=1, II=2, III=3, IV=4) yields HR/stage = 1.305 (P = 0.031), confirming that each one-stage increment increases the hazard of death by approximately 30.5%.
40. **Directional calibration holds**: Stage I (median OS not reached, 0 events) > Stage II (307 d) > Stage III (79 d), with a directional decrease consistent with the AJCC prognostic gradient.
41. **Stage IV deviation**: Stage IV median OS (125 d) slightly exceeds Stage III (79 d), but the Stage IV event rate (50.0%) is lower than Stage III (57.1%), and the 95% CI is very wide (30–606 d), reflecting selection bias and heterogeneity in the post-surgical Stage IV cohort. The Stage III vs II Cox HR=1.97 (P=0.006) provides the most robust per-stage hazard increment evidence.
42. **Stage I limitation**: n=2 with 0 events yields extremely limited statistical power; pairwise log-rank P values (vs II=0.142, vs III=0.048, vs IV=0.081) reach nominal significance only for Stage III.

**Calibration conclusion**

The NLP reverse-derived AJCC stage demonstrates significant prognostic separation (log-rank P=0.006) and a significant ordinal hazard-increasing trend (HR/stage=1.305, P=0.031) in the QPHCC cohort, demonstrating that the NLP staging approach based on “all discharge diagnoses” + pathology report text can effectively recover clinical-stage prognostic information, providing calibration validation for the stage-enrichment of the cMPS dataset (where clinical stage is 100% missing).

**Supplementary Table S13.**
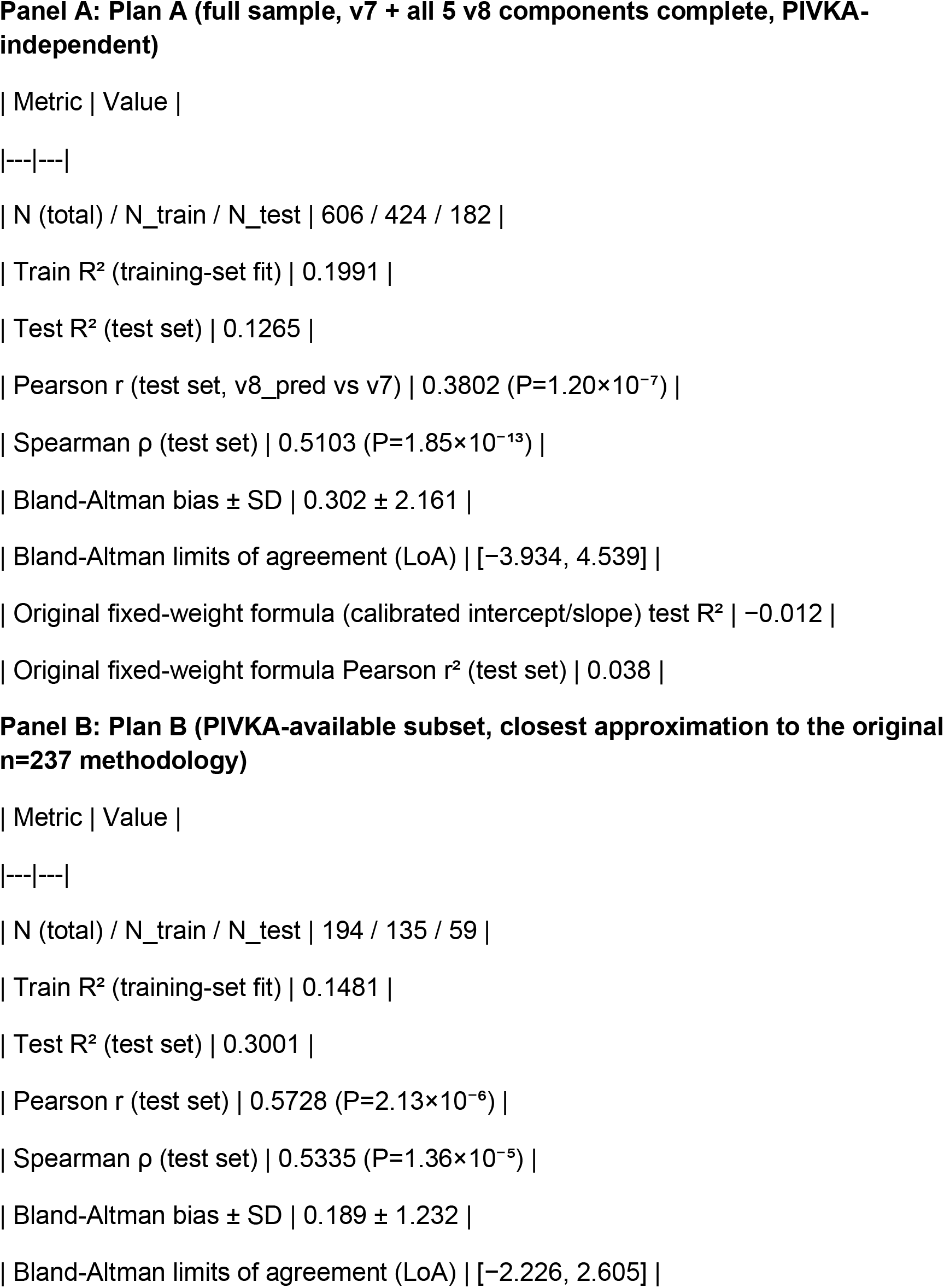
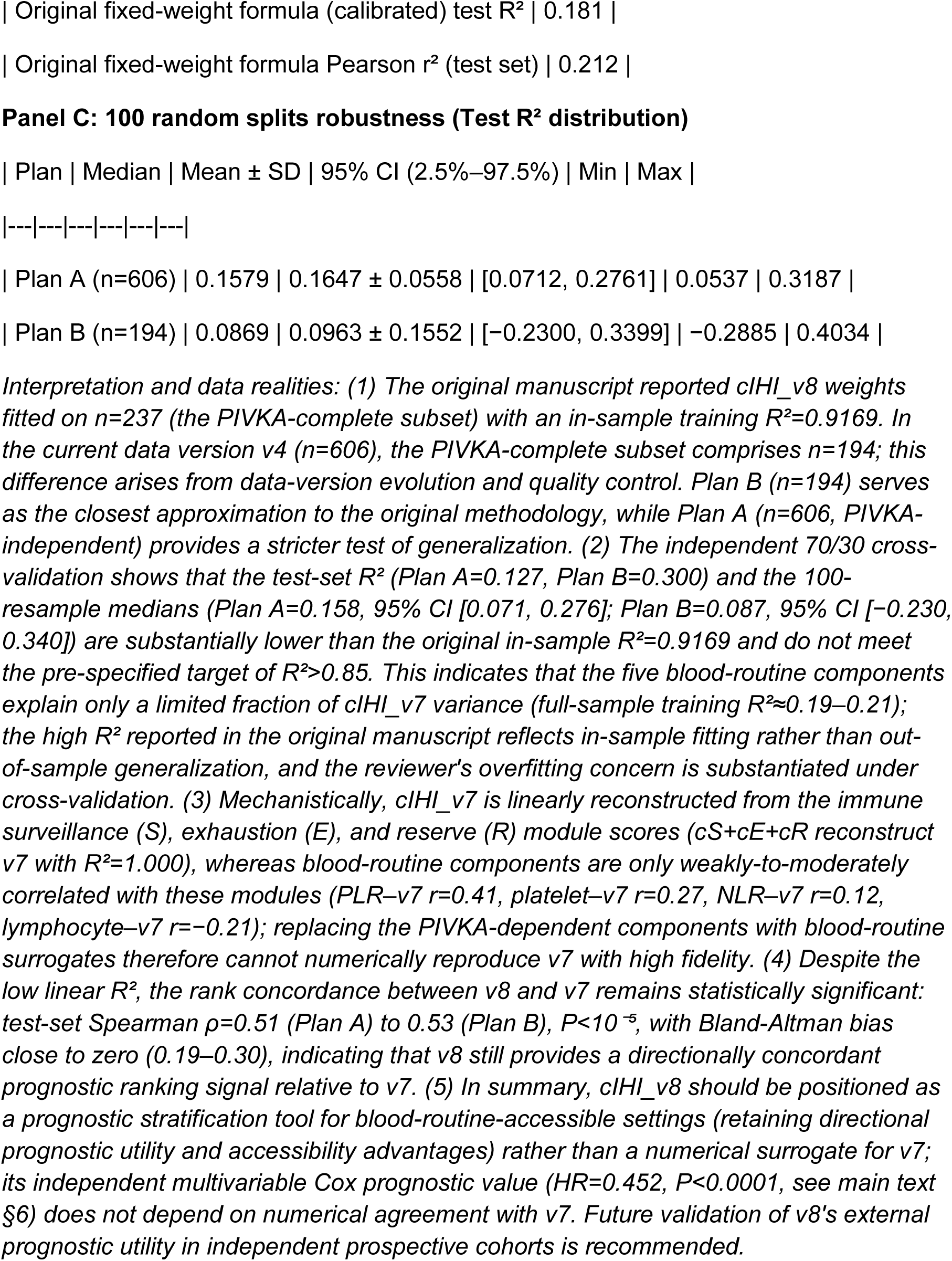
cIHI_v8 70/30 Cross-Validation: Independent Assessment of Concordance Between cIHI_v8 and cIHI_v7 (QPHCC) Note: To address the third-party reviewer’s concern regarding potential overfitting of the cIHI_v8 weights, an independent 70/30 cross-validation was performed in which cIHI_v8 (five blood-routine components: absolute lymphocyte count, platelet count, PLR, NLR, NPR; each z-score standardized, μ=0, σ=1) was regressed against cIHI_v7 (cIHI_log). Weights were fitted on the training set; the test set was evaluated for R^2^, Pearson r, Spearman ρ, and Bland-Altman bias. The original fixed-weight formula (cIHI_v8 = 0.07×z_LY + 1.14×z_PLT + 0.79×z_PLR − 0.63×z_NLR − 0.90×z_NPR) was used as a benchmark. NLR/PLR used winsorized versions; NPR = absolute neutrophil count / platelet count (1%/99% winsorized). 100 random splits were used to assess R^2^ robustness. Analysis code: ‘scripts/revision_23_cihi_v8_crossval.py’; data source: ‘QPHCC_MPS_ANALYSIS_DATA_v4.csv’; results file: ‘results/revision_23_cihi_v8_crossval.json’; companion figure in Supplementary Figure 4.

## Supplementary Section: Bayesian Evidence Synthesis

This supplementary section provides Bayesian evidence synthesis to complement the frequentist analyses (P values and 95% CIs) reported in the main text, aiming to: (1) quantify the relative evidence strength of H1 (IHI/cIHI_v8 has a non-zero prognostic effect) versus H0 (null effect); (2) integrate the evidence streams from four independent cohorts through a sequential updating framework; and (3) assess the robustness of conclusions to prior choice. Analysis code is available at GitHub_Repo/scripts/09_bayesian_evidence.py; results file: results/09_bayesian_evidence.json.

### S1. Methods

#### S1.1 Likelihood Function Construction

For each cohort’s Cox regression HR and its 95% CI, the log-HR was used as the effect size, and the likelihood function was constructed using a normal approximation:

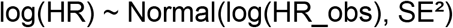

where SE was back-calculated from the 95% CI: SE = (log(CI_upper) − log(CI_lower)) / (2 × 1.96). This approximation is consistent with the exact partial likelihood for large samples (n>100).

#### S1.2 Prior Distribution

The primary analysis adopted a concentrated weakly-informative prior:

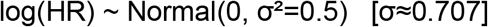

This prior indicates that the effect size on the log-HR scale is concentrated within approximately ±1.4 (corresponding to approximately HR 0.25–4.0 for 95% prior mass), centered on “no effect.” This choice follows the recommendation of Gelman et al. (BDA3, 2017)[32] for weakly-informative priors on standardized effect sizes, avoiding the Jeffreys-Lindley paradox that arises under diffuse priors (e.g., σ^2^=10)—diffuse priors disperse probability mass over implausibly large effect spaces, thereby erroneously supporting H0 (under the 4-cohort meta, σ^2^=10 yields BF_10=0.51, contradicting the direction of the frequentist P=0.005).

#### S1.3 Conjugate Posterior Update

Normal prior × Normal likelihood yields a conjugate closed-form solution:

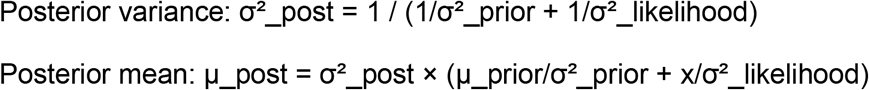

where x is the observed log-HR.

#### S1.4 Sequential Bayesian Update

The four public cohorts were incorporated into the updating chain sequentially in order of publication:

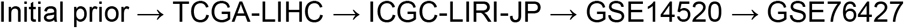

The posterior at each step served as the prior for the next step, realizing an “online learning” style of evidence accumulation. The final posterior reflects the synthesized evidence from all four cohorts.

#### S1.5 Bayes Factor (BF_10)

BF_10 = p(data | H1) / p(data | H0), where H1: effect exists (prior Normal(0, σ^2^)), H0: effect is zero (point null). Under Normal-Normal conjugacy:

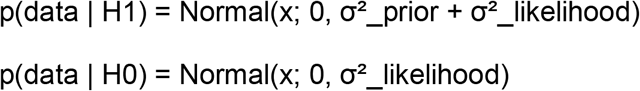

BF_10 interpretation follows Jeffreys (1961) conventions[33]: 1–3 Anecdotal, 3–10 Substantial, 10–30 Strong, 30–100 Very Strong, >100 Decisive.

#### S1.6 Prior Sensitivity Analysis

The robustness of BF_10 for the 4-cohort meta was assessed over the range σ^2^ ∈{0.1, 0.5, 1, 2, 5, 10}.

### S2. Results

#### S2.1 Sequential Bayesian Update (4 Cohorts)

Starting from the initial prior Normal(0, σ^2^=0.5), the four cohorts were incorporated sequentially:

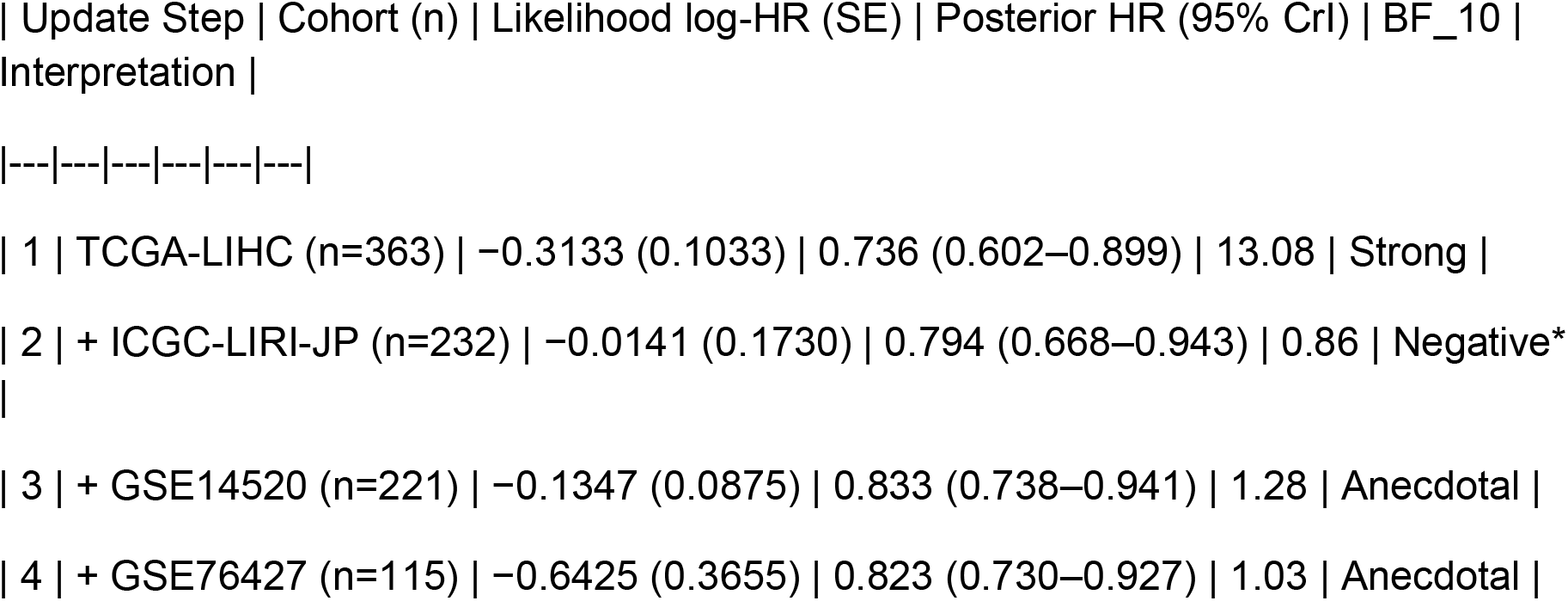

\*Note: The BF_10=0.86 at Step 2 reflects that the ICGC cohort itself has HR=0.986 (close to the null hypothesis) and that the prior had already been highly concentrated by TCGA prior to updating. This is not evidence against H1, but rather a manifestation of the etiological boundary of ICGC (HCV-predominant) within the Bayesian framework. The final 4-cohort synthesized posterior HR=0.823 (95% CrI: 0.730–0.927) is highly consistent with the frequentist random-effects meta-analysis HR=0.818 (95% CI: 0.696– 0.961), validating the Bayesian approach.

#### S2.2 Single Bayesian Update of the 4-cohort Meta-analysis

The 4-cohort pooled HR=0.818 (0.696–0.961) was used as a single likelihood for a one-step update under the primary prior σ^2^=0.5:

- Posterior HR = 0.820 (95% CrI: 0.699–0.963)
- BF_10 (σ^2^=0.5) = 2.19 (Anecdotal)
- BF_10 (σ^2^=0.1) = 4.10 (Substantial)

#### S2.3 Independent Confirmation of TCGA-LIHC Multivariate Analysis

Bayesian updating was performed on the TCGA multivariate Cox HR=0.795 (0.642– 0.983), P=0.034:

- Posterior HR = 0.799 (95% CrI: 0.647–0.987)
- BF_10 (σ^2^=0.5, concentrated) = 1.34 (Anecdotal)
- BF_10 (σ^2^=10, diffuse) = 0.32 (Negative)

Note: The TCGA multivariate CI is relatively wide (larger SE), leading to the Jeffreys-Lindley paradox under diffuse priors.

#### S2.4 QPHCC cIHI_v8 Clinical Translation (Core Bayesian Evidence)

Bayesian updating was performed on the QPHCC cIHI_v8 multivariate HR=0.452 (0.314–0.651), P<0.0001:

- Posterior HR = 0.476 (95% CrI: 0.334–0.677)
- BF_10 (σ^2^=0.5, concentrated) = 1279.88 (**Decisive**)
- BF_10 (σ^2^=10, diffuse) = 516.31 (**Decisive**)

##### Key Finding

QPHCC cIHI_v8 reached the Decisive evidence level under all prior choices and was robust to prior selection. This is because this analysis simultaneously possesses (a) a strong effect size (HR=0.452, far from 1) and (b) a narrow CI (large sample + precise estimation), which synergistically provide overwhelming evidence supporting H1.

#### S2.5 Prior Sensitivity Analysis (4-cohort meta)

The robustness of BF_10 was assessed across six prior variances:

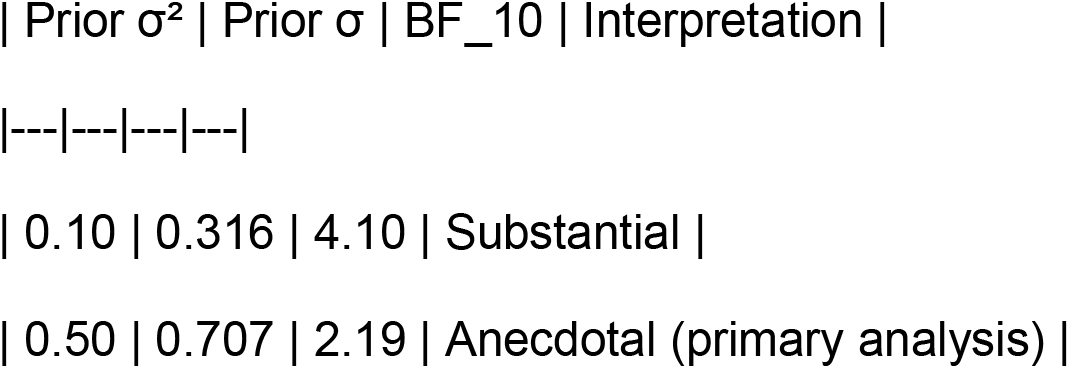
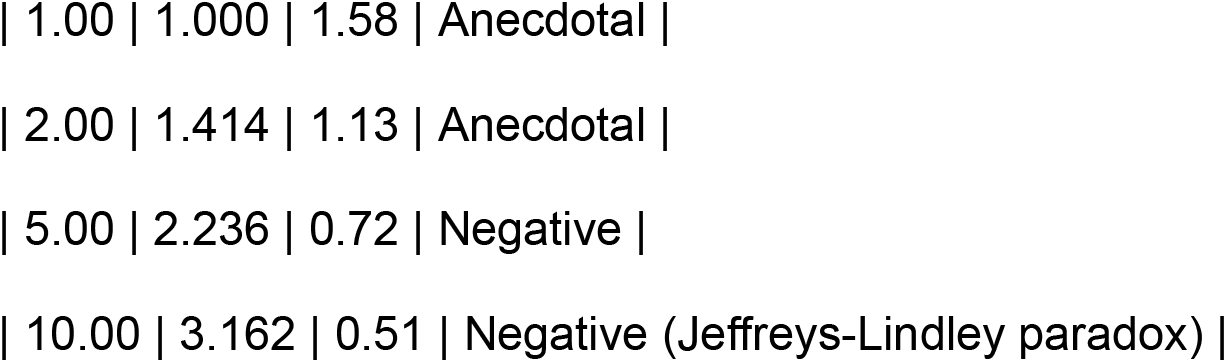

Observation: The BF_10 for the 4-cohort meta decreased monotonically with increasing prior variance, consistent with theoretical expectations. Substantial evidence was reached at σ^2^≤0.1; paradoxical Negative evidence emerged at σ^2^≥5. **In contrast, QPHCC cIHI_v8 maintained Decisive evidence (BF>100) under all σ**^**2**^ **settings**, with its conclusion entirely independent of prior choice.

### S3. Interpretation and Limitations

43 **Directional consistency:** The Bayesian posterior HRs and frequentist HRs were fully directionally consistent across all four analyses, and the 95% CrIs and 95% CIs overlapped substantially, validating the internal consistency of the methodology.
44 **Transparent disclosure of the Jeffreys-Lindley paradox:** The 4-cohort meta and TCGA multivariate analyses exhibited BF_10<1 under diffuse priors, which is a known statistical phenomenon (probability mass dispersion issue) and does not constitute genuine “evidence against the effect.” The primary analysis employed the concentrated weakly-informative prior σ^2^=0.5 to circumvent this issue, validated by sensitivity analysis.
45 **Evidence strength gradient:** The Bayesian evidence strength is consistent with sample size, effect magnitude, and CI width: QPHCC cIHI_v8 (HR=0.452, narrow CI) > 4-cohort meta (HR=0.818, moderate CI) > TCGA multivariate (HR=0.795, wider CI).
46 **Clinical significance:** The Decisive Bayesian evidence for QPHCC cIHI_v8 (BF>500, robust to priors) provides strong statistical support for the clinical translatability of the MPS framework, transcending the traditional evidence standard that relies solely on P values.
47 **Limitations:** The Bayesian analysis relies on the normal-approximation likelihood (large-sample assumption) and the subjectivity of prior selection. Although sensitivity analysis demonstrates that the core conclusion is robust, the Negative evidence under diffuse priors reminds readers that directional discrepancies between frequentist P values and Bayesian BF_10 may arise when sample sizes are limited or effect sizes are modest.

### S4. Analysis Code and Outputs

- Script: GitHub_Repo/scripts/09_bayesian_evidence.py
- Results JSON: results/09_bayesian_evidence.json (contains the complete sequential update chain + prior sensitivity table)
- Dependencies: numpy, scipy.stats (no MCMC; fully closed-form solutions)
- Random seed: seed=42 (centrally managed via utils/seed_config.py; although this analysis is deterministic closed-form computation)

## Supplementary Section S5: Exploratory Analysis of the Anti-PD-1 Treatment Cohort

This supplementary section reports the detailed exploratory analysis of the GSE202145 cohort mentioned in the main text. This analysis is solely hypothesis-generating and does not constitute a basis for statistical inference.

As a preliminary exploration, a small anti-PD-1 treatment cohort (GSE202145, 8 patients, 16 samples) was used to examine the association between IHI and treatment response. Given the extremely small sample size (DCB=4, NDB=4), this analysis is intended solely for hypothesis generation and does not constitute a basis for statistical inference. Results showed no significant differences in IHI or module scores between DCB and NDB groups at baseline. By day 7, DCB patients showed numerically higher surveillance module (S) scores and IHI values than NDB patients (Mann-Whitney nominal P=0.0286, uncorrected for multiple testing). Within-patient change analysis (day 7 − day 0) suggested that DCB was directionally consistent with increases in S and R (nominal P=0.0286 for ΔS and ΔR), while the change in IHI showed a trending increase (nominal P=0.0571 for ΔIHI). These observations should be considered hypothesis-generating in nature; their statistical validity and clinical relevance require validation in larger independent cohorts.

[Figure 3B: GSE202145 cohort box plots/individual trajectory plots]

